# Multiomic Body Mass Index signatures in blood reveal clinically relevant population heterogeneity and variable responses to a healthy lifestyle intervention

**DOI:** 10.1101/2022.01.20.22269601

**Authors:** Kengo Watanabe, Tomasz Wilmanski, Christian Diener, John C. Earls, Anat Zimmer, Briana Lincoln, Jennifer J. Hadlock, Jennifer C. Lovejoy, Sean M. Gibbons, Andrew T. Magis, Leroy Hood, Nathan D. Price, Noa Rappaport

**Affiliations:** Institute for Systems Biology, Seattle, WA 98109, USA; Thorne HealthTech, New York, NY 10019, USA; Department of Bioengineering, University of Washington, Seattle, WA 98195, USA; eScience Institute, University of Washington, Seattle, WA 98195, USA; Phenome Health, Seattle, WA 98109, USA

## Abstract

Multiomic profiling can reveal population heterogeneity for both health and disease states. Obesity drives a myriad of metabolic perturbations in individuals and is a risk factor for multiple chronic diseases. Here, we report a global atlas of cross-sectional and longitudinal changes in 1,111 blood analytes associated with variation in Body Mass Index (BMI), as well as the multiomic associations with host polygenic risk scores and gut microbiome composition, from a cohort of 1,277 individuals enrolled in a wellness program. Machine learning model predictions of BMI from blood multiomics captured heterogeneous phenotypic states of host metabolism and gut microbiome composition, better than classically-measured BMI. Moreover, longitudinal analyses identified variable BMI trajectories for different omics measures in response to a healthy lifestyle intervention; metabolomics-inferred BMI decreased to a greater extent than actual BMI, while proteomics-inferred BMI exhibited greater resistance to change. Our analyses further revealed blood analyte–analyte associations that were significantly modified by metabolomics-inferred BMI and partially reversed in the metabolically obese population during the intervention. Taken together, our findings provide a blood atlas of the molecular perturbations associated with changes in obesity status, serving as a valuable resource to robustly quantify metabolic health for predictive and preventive medicine.

## Introduction

Obesity has been increasing in prevalence over the past four decades in adults, adolescents, and children around most of the world^1,2^. Many studies have demonstrated that obesity is a major risk factor for multiple chronic diseases such as type 2 diabetes mellitus (T2DM), metabolic syndrome (MetS), cardiovascular disease (CVD), and certain types of cancer^3–6^. In individuals with obesity, even a 5% loss in body weight can improve metabolic and cardiovascular health^7^, and weight loss through lifestyle interventions can reduce the risk for obesity-related chronic diseases^8^. Nevertheless, obesity and its physiological manifestations can vary widely across individuals, necessitating additional research to better understand this prevalent health condition.

Most commonly, obesity is quantified using the anthropometric Body Mass Index (BMI), defined as the body weight divided by body height squared [kg m^−2^]. While BMI does not directly measure body composition, BMI correlates well at the population level with direct measurements of body fat percentage using computed tomography (CT), magnetic resonance imaging (MRI), or dual-energy X-ray absorptiometry (DXA) (partial Pearson’s *r* = 0.74–0.84)^9^. As an easily calculated and commonly understood measure among researchers, clinicians, and the general public, BMI is widely used for the primary diagnosis of obesity, and changes in BMI are often used to assess the efficacy of lifestyle interventions.

At the same time, there are considerable limitations to BMI as a surrogate measure of health state; e.g., differences in body composition can lead to misclassification of people with a high muscle-to-fat ratio (e.g., athletes) as the individual with obesity, and can undervalue metabolic improvements in health following exercise^10^. A meta-analysis showed that the common obesity diagnoses based on BMI cutoffs had high specificity but low sensitivity in identifying individuals with excess body fat^11^. The misclassification is likely due, in part, to the differences in BMI thresholds for obesity across different ethnic populations^12^, as well as the existence of a metabolically unhealthy, normal-weight (MUNW) group within the normal BMI class^13,14^. Likewise, there are health-heterogeneous groups among the individuals with obesity: metabolically healthy obese (MHO) and metabolically unhealthy obese (MUO). While most individuals in the MHO group are not necessarily healthy but simply healthier than individuals in the MUO group^15^, the transition from MHO to MUO phenotype may be a preceding step to the development of obesity-related chronic diseases^16^. Moreover, this transition is potentially preventable through lifestyle interventions^17^. Altogether, BMI is unequivocally useful at the population level, but too crude to capture a variety of heterogeneous metabolic health states.

Recent omics studies have demonstrated how blood omic profiles contain information relevant to a wide range of human health conditions; e.g., blood proteomics captured 11 health indicators such as the liver fat measured by ultrasound and the body composition measured by DXA^18^, while blood metabolomics tended to reflect dietary intake, lifestyle patterns, and gut microbiome profiles^19,20^. Intriguingly, a machine learning model that was trained to predict BMI using 49 BMI-associated blood metabolites captured obesity-related clinical measurements (e.g., insulin resistance, visceral fat percentage) better than observed BMI or genetic predisposition for high BMI^21^. Moreover, in a recent study on coronary artery disease, another blood metabolomics-based model of BMI efficiently reflected differences between individuals with or without acute coronary syndrome (ACS)^22^. Thus, while a single targeted metric (e.g., body composition) or a specific biomarker (e.g., leptin, adiponectin^23^) provides useful information, multiomic blood profiling has the potential to comprehensively bridge the multifaceted gaps between BMI and heterogeneous physiological states.

In this study, we report heterogeneous molecular signatures of obesity by leveraging a cohort of 1,277 individuals with a wealth of phenotypic data, including human genomes and longitudinal measurements of metabolomics, proteomics, clinical laboratory tests, gut microbiomes, physical activity (i.e., wearables), and health/lifestyle questionnaires, and by employing machine learning to predict BMI. Blood-based analytes across all studied omics platforms exhibit strong performance in predicting measured BMI, explaining 48–78% of the variance in our main study cohort. We further show that multiomic phenotyping captures more refined levels of heterogeneity in metabolic states accompanying obesity, which is not apparent when using measured BMI. Moreover, longitudinal analyses demonstrate variable changes in blood analytes across the studied omics platforms during a healthy lifestyle intervention; i.e., plasma metabolomics exhibited a stronger response to the intervention than measured BMI, while plasma proteomics exhibited a weaker response within a one-year span. Our findings highlight the utility and translational potential of blood multiomic profiling for investigating the complex molecular phenotypes underlying obesity and weight loss.

## Results

### Plasma multiomics captured 48–78% of the variance in BMI

To investigate the molecular phenotypic perturbations associated with obesity, we selected a study cohort of 1,277 adults who participated in a scientific wellness program (Arivale)^20,24–29^ and whose datasets included coupled measurements of plasma metabolomics, proteomics, and clinical laboratory tests from the same blood draw (Fig. 1a; see Methods). This study design allowed us to directly investigate the similarities and differences between omics platforms with regards to how they reflected the physiological health state of each individual across the BMI spectrum. This cohort was characteristically female (64.3%), middle-aged (mean ± s.d.: 46.6 ± 10.8 years), and white (69.7%) (Supplementary Fig. 1a–c, Supplementary Data 1). Based on the World Health Organization (WHO) international standards for BMI cutoffs (underweight: <18.5 kg m^−2^, normal: 18.5–25 kg m^−2^, overweight: 25–30 kg m^−2^, obese: ≥30 kg m^−2^)^12^, the baseline BMI prevalence was similar among normal, overweight, and obese classes, while only 0.8% of participants were in the underweight class (underweight: 10 participants (0.8%), normal: 426 participants (33.4%), overweight: 391 participants (30.6%), obese: 450 participants (35.2%)).

**Figure 1.**
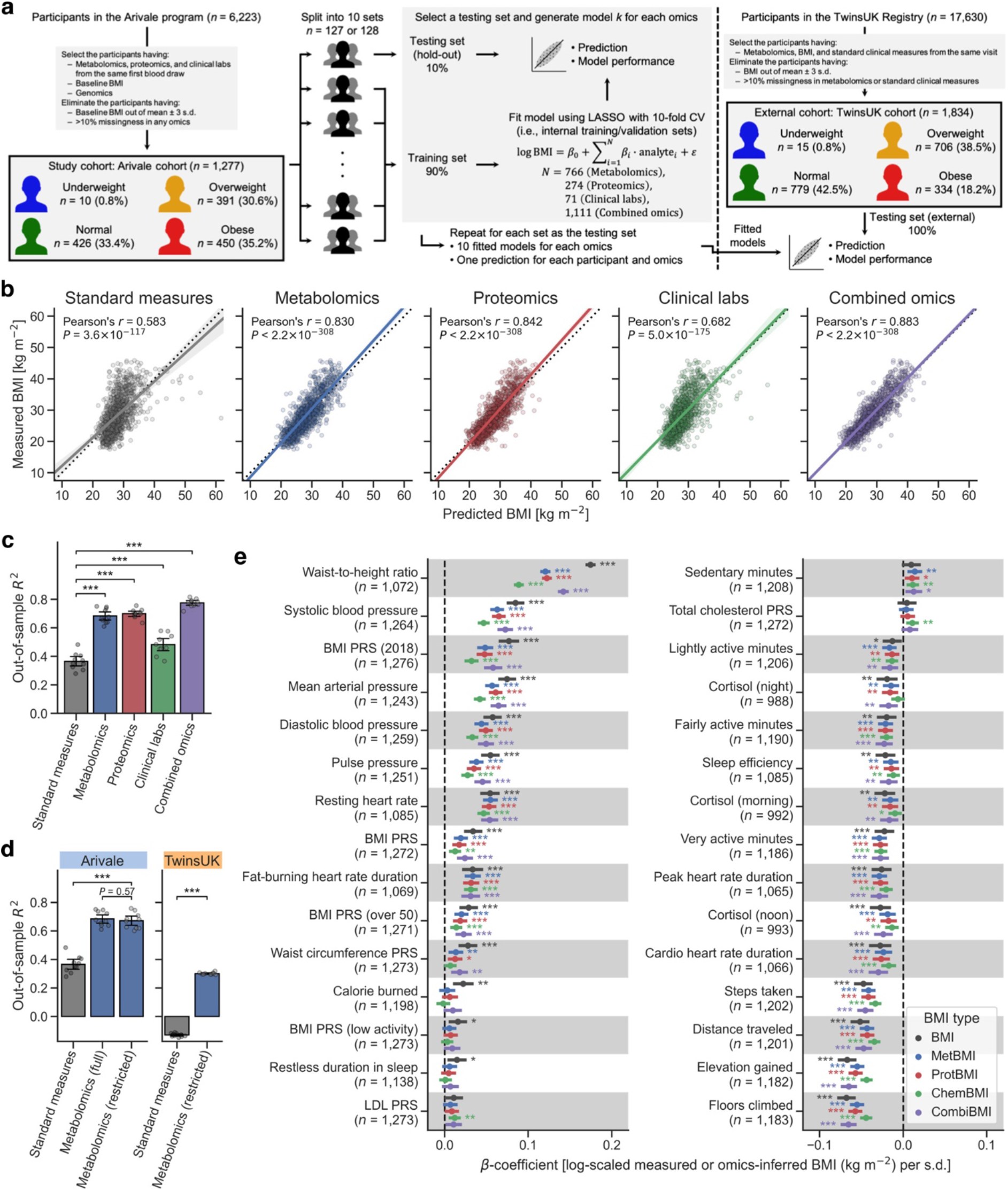
Plasma multiomics captured 48–78% of the variance in BMI. **a** Overview of study cohorts and the omics-based Body Mass Index (BMI) model generation. LASSO: least absolute shrinkage and selection operator, CV: cross-validation. **b** Correlation between the measured and predicted BMIs. The solid line is the ordinary least squares (OLS) linear regression line with 95% confidence interval (CI), and the dotted line is measured BMI = predicted BMI. Standard measures: OLS linear regression model with sex, age, triglycerides, high-density lipoprotein (HDL)-cholesterol, low-density lipoprotein (LDL)-cholesterol, glucose, insulin, and homeostatic model assessment for insulin resistance (HOMA-IR) as regressors; *P*: adjusted *P*-value of two-sided Pearson’s correlation test with the Benjamini–Hochberg method across the five categories. *n* = 1,277 participants. **c, d** Model performance of each fitted BMI model. Out-of-sample *R*^2^ was calculated from each corresponding hold-out testing set (**c**, Arivale in **d**) or from the external testing set (TwinsUK in **d**). Metabolomics (full): LASSO model trained by all 766 metabolites of the Arivale dataset, Metabolomics (restricted): LASSO model trained by the common 489 metabolites in the Arivale and TwinsUK datasets (see Supplementary Fig. 3). Note that Standard measures and Metabolomics (full) of Arivale in **d** are the same with corresponding ones in **c**. Data: mean with 95% CI, *n* = 10 models. ***Adjusted *P* < 0.001 in two-sided Welch’s *t*-test with the Benjamini–Hochberg method across the four (**c**) or three (**d**) comparisons. **e** Association between omics-inferred BMI and physiological feature. For each of the 51 numeric physiological features (Supplementary Data 4), *β*-coefficient was estimated using OLS linear regression model with the measured or omics-inferred BMI as dependent variable and sex, age, and ancestry principal components (PCs) as covariates. Presented are the 30 features that were significantly associated with at least one of the BMI types after multiple testing adjustment with the Benjamini–Hochberg method across the 255 (51 features × 5 BMI types) regressions. BMI: measured BMI, MetBMI: metabolomics-inferred BMI, ProtBMI: proteomics-inferred BMI, ChemBMI: clinical chemistries-inferred BMI, CombiBMI: combined omics-inferred BMI, PRS: polygenic risk score, *n*: the number of assessed participants. Data: estimate with 95% CI. *Adjusted *P* < 0.05, **adjusted *P* < 0.01, ***adjusted *P* < 0.001.

Leveraging the baseline measurements of plasma molecular analytes (766 metabolites, 274 proteins, and 71 clinical laboratory tests; Supplementary Data 2), we trained machine learning models to predict baseline BMI (i.e., not forecast a future outcome but calculate an out-of-sample outcome) for each of the omics platforms (metabolomics, proteomics, and clinical labs) or in combination (combined omics of all metabolomics, proteomics, and clinical labs): metabolomics-based, proteomics-based, clinical labs (chemistries)-based, and combined omics-based BMI (MetBMI, ProtBMI, ChemBMI, and CombiBMI, respectively) models. To address multicollinearity among the analytes (Supplementary Fig. 2a) and to obtain predictions for all participants, we applied a tenfold iteration scheme of the least absolute shrinkage and selection operator (LASSO) algorithm with tenfold cross-validation (CV) (Fig. 1a; see Methods). This approach generated ten fitted sparse models for each omics category (Supplementary Data 3) and one single testing (hold-out) set-derived prediction from each omics category for each participant. The resulting models retained (i.e., assigned non-zero *β*-coefficient to) 62 metabolites, 30 proteins, 20 clinical laboratory tests, and 132 analytes across all ten MetBMI, ProtBMI, ChemBMI, and CombiBMI models, respectively, which exhibited low collinearity (Supplementary Fig. 2b, c) as expected from the LASSO algorithm^30^. In contrast to a model including obesity-related standard clinical measures (i.e., ordinary least squares (OLS) linear regression model with sex, age, triglycerides, high-density lipoprotein (HDL)-cholesterol, low-density lipoprotein (LDL)-cholesterol, glucose, insulin, and homeostatic model assessment for insulin resistance (HOMA-IR) as regressors; StandBMI model), each omics-based model demonstrated significantly higher performance in BMI prediction, ranging from out-of-sample *R*^2^ = 0.48 (ChemBMI) to 0.70 (ProtBMI) compared to 0.37 (StandBMI) (Fig. 1b, c). The CombiBMI model exhibited the best performance in BMI prediction (out-of-sample *R*^2^ = 0.78; Fig. 1c), but the variances explained were not completely additive, suggesting that, although there is a considerable overlap in the signal detected by each omics platform, different omic measurements still contain non-redundant information regarding BMI. Additionally, these results were consistent in sex-stratified models, with the exception of male ChemBMI model that tended to exhibit higher performance than StandBMI model without statistical significance (Supplementary Fig. 2d).

To confirm the generalizability of our results, we investigated an external cohort of 1,834 adults from the TwinsUK registry^31^, whose datasets included serum metabolomics^32^ and the aforementioned standard clinical measures (Fig. 1a; see Methods). This external cohort was demographically distinct from the Arivale cohort (Supplementary Fig. 1d–f, Supplementary Data 1); the TwinsUK cohort was overwhelmingly female (96.7%), senior (mean ± s.d.: 61.4 ± 9.0 years), and white (99.2%), and consisted of 15 (0.8%), 779 (42.5%), 706 (38.5%), and 334 (18.2%) participants in the underweight, normal, overweight, and obese BMI classes, respectively. To manage the differences in the metabolomics panels, we regenerated MetBMI models in the Arivale cohort, while restricting the metabolomic features to an overlapping set of 489 metabolites between the Arivale and TwinsUK panels (called restricted model). Although 25 of the retained metabolites in the original MetBMI models were replaced with other metabolites due to their absences in the restricted panel, 35 of the remaining 37 metabolites were consistently retained across the restricted MetBMI models (Supplementary Fig. 3a). Moreover, *β*-coefficients for the retained metabolites and MetBMI predictions for the Arivale cohort were consistent between the original and restricted models (Supplementary Fig. 3b, c). We then calculated BMI predictions for the TwinsUK cohort using the StandBMI and restricted MetBMI models that were fitted to the Arivale datasets. The restricted MetBMI model exhibited a lower absolute performance on the TwinsUK cohort compared to the Arivale cohort, but a significantly higher performance than StandBMI model (out-of-sample *R*^2^ = 0.30 (MetBMI), −0.13 (StandBMI); Fig. 1d, Supplementary Fig. 3d), confirming that blood metabolomics generally captures BMI better than the standard clinical measures.

BMI has been reported to be associated with multiple anthropometric and clinical measures, such as waist circumference (WC), blood pressure, sleep quality, and several polygenic risk scores (PRSs)^3,4,15,27,33^. Thus, we examined the association between the omics-inferred BMI and each of the available numeric physiological measures (see Methods; Supplementary Data 4). Among the 51 assessed features, measured BMI was significantly associated with 27 features (false discovery rate (FDR) < 0.05) including daily physical activity measures from wearable devices, waist-to-height ratio (WHtR), blood pressure, and BMI PRS (Fig. 1e). With minor differences in effect sizes, these BMI-associated features were concordantly associated with each omics-inferred BMI (Fig. 1e), indicating that the omics-inferred BMIs primarily maintain the characteristics of classical BMI in terms of anthropometric, genetic, lifestyle, and physiological associations.

### Omics-based BMI estimates captured the variation in BMI better than any single analyte

Because our LASSO linear regression model showed comparable performance to elastic net (EN) and ridge linear regression models and a non-linear random forest (RF) regression model (Supplementary Fig. 4a, b) and because LASSO model *β*-coefficients are generally easier to be interpreted, we chose to focus on the LASSO models. However, the LASSO algorithm randomly retains variables from highly collinear groups, and sets *β*-coefficients of the other variables to zero. To confirm the robustness of the variable selection process, we iterated the LASSO modeling while removing the strongest analyte (i.e., the analyte that had the highest absolute value for the mean of the ten *β*-coefficients) from the input omic dataset at the end of each iteration. If a variable is indispensable for a model, the performance should largely decrease after removing it. In all omics categories, a steep decay in the out-of-sample *R*^2^ was observed in the first 5–9 iterations (Supplementary Fig. 2e–h), suggesting that, at least, the top 5–9 variables that had the highest absolute *β*-coefficient values in the original LASSO models were indispensable for predicting BMI. Interestingly, the overall slope of *R*^2^ in MetBMI model decayed more gradually compared to ProtBMI and ChemBMI models (Supplementary Fig. 2e–g), implying that metabolomics data contain more redundant information about BMI than the other omics data. Although larger number of metabolites in the input dataset might be a plausible explanation, the proportion of the variables that were robustly retained across all ten LASSO models (Supplementary Fig. 5) to the variables that were retained in at least one of the ten LASSO models was lower in MetBMI model compared to ProtBMI and ChemBMI models (MetBMI: 62/209 metabolites ≈ 30%, ProtBMI: 30/74 proteins ≈ 41%, ChemBMI: 20/41 clinical laboratory tests ≈ 49%), confirming the higher level of redundancy within metabolomics data. Nevertheless, metabolites still constituted 58% of the 132 analytes that were retained across all ten CombiBMI models (77 metabolites, 51 proteins, 4 clinical laboratory tests; Fig. 2a), suggesting that each of the omics categories possesses unique information about BMI. The strongest predictors in CombiBMI model were primarily proteins; e.g., analytes having the mean absolute *β*-coefficient > 0.02 (i.e., affecting more than ∼2% BMI in prediction per 1 s.d. of its change, according to the Taylor/Maclaurin series: e^*β*^ ≈ 1 + *β* when *β* << 1) were leptin (LEP), adrenomedullin (ADM), and fatty acid-binding protein 4 (FABP4) as the positive predictors and insulin-like growth factor-binding protein 1 (IGFBP1) and advanced glycosylation end-product specific receptor (AGER; also described as receptor of AGE, RAGE) as the negative predictors. Note that these strongest proteins were consistent in the EN models (Supplementary Fig. 4c–f) and had high importance in the ridge and RF models (Supplementary Fig. 4g, h).

**Figure 2.**
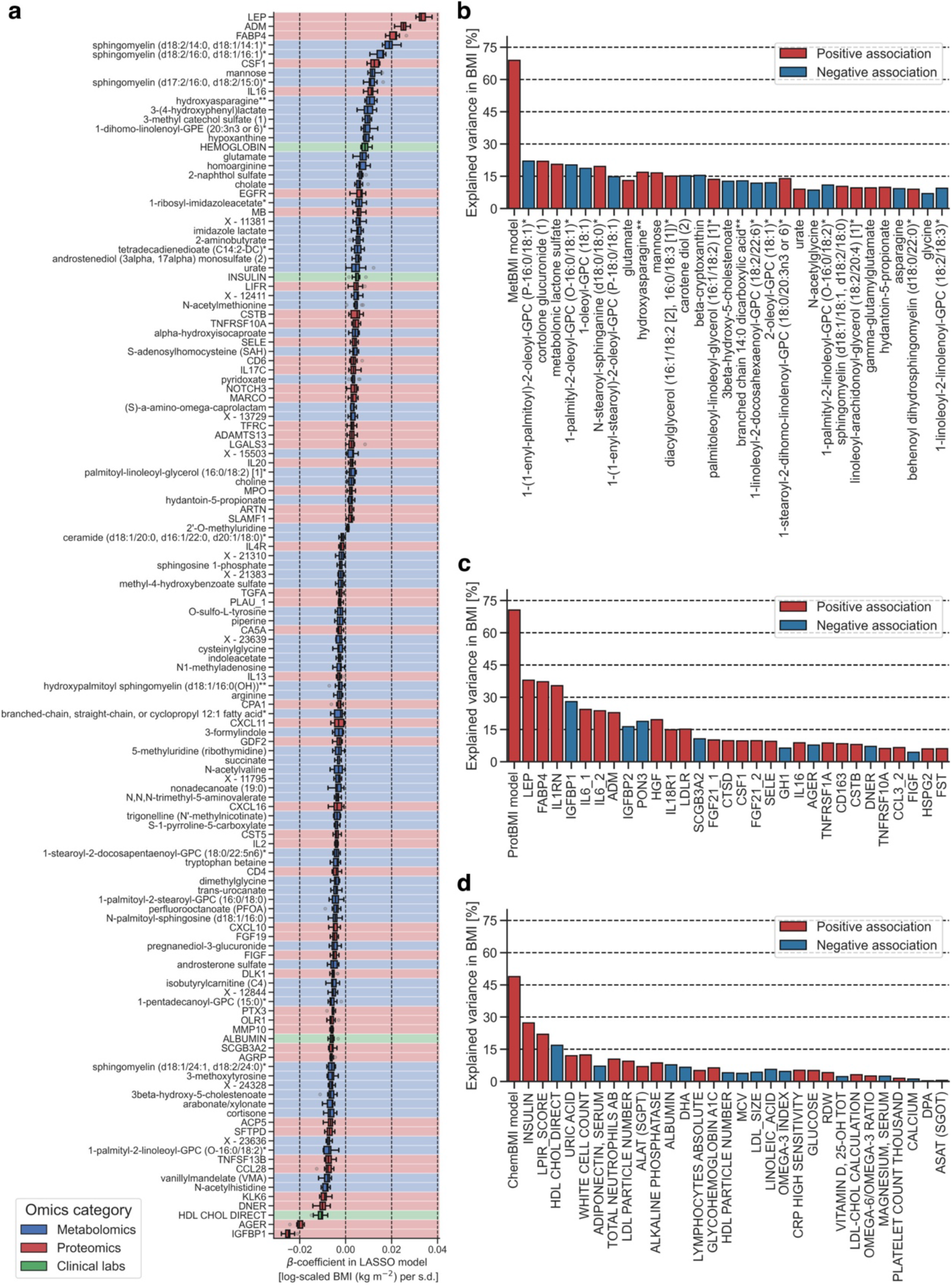
Omics-based BMI estimates captured the variance in BMI better than any single analyte. **a** The variables that were retained across all ten combined omics-based Body Mass Index (CombiBMI) models (132 analytes: 77 metabolites, 51 proteins, and 4 clinical laboratory tests). *β*-coefficient was obtained from the fitted CombiBMI model with least absolute shrinkage and selection operator (LASSO) regression. Each background color corresponds to the analyte category. Data: median (center line), [*Q*_1_, *Q*_3_] (box limits), [*x*_min_, *x*_max_] (whiskers), where *Q*_1_ and *Q*_3_ are the 1st and 3rd quartile values, and *x*_min_ and *x*_max_ are the minimum and maximum values in [*Q*_1_ − 1.5 × IQR, *Q*_3_ + 1.5 × IQR] (IQR: the interquartile range, *Q*_3_ − *Q*_1_), respectively; *n* = 10 models. **b**–**d** Univariate explained variance in BMI by each metabolite (**b**), protein (**c**), or clinical laboratory test (**d**). BMI was independently regressed on each of the analytes that were retained in at least one of the ten LASSO models (209 metabolites, 74 proteins, 41 clinical laboratory tests; Supplementary Data 5), using ordinary least squares (OLS) linear regression with sex, age, and ancestry principal components (PCs) as covariates. Multiple testing was adjusted with the Benjamini–Hochberg method across the 210 (**b**), 75 (**c**), or 42 (**d**) regressions, including each omics-based BMI (MetBMI: metabolomics-based BMI, ProtBMI: proteomics-based BMI, ChemBMI: clinical chemistries-based BMI) model as reference. Among the analytes that were significantly associated with BMI (180 metabolites, 63 proteins, 30 clinical laboratory tests), only the top 30 significant analytes are presented with their univariate variances.

At the same time, the existence of these strong and consistently-retained predictors in the omics-based BMI models implied that a single analyte might be a suitable biomarker to predict BMI. To address this possibility, we regressed BMI independently on each of the analytes that were retained in at least one of the ten LASSO models (MetBMI: 209 metabolites, ProtBMI: 74 proteins, ChemBMI: 41 clinical laboratory tests; Supplementary Data 5). Among the analytes that were significantly associated with BMI (180 metabolites, 63 proteins, 30 clinical laboratory tests), only LEP, FABP4, and interleukin 1 receptor antagonist (IL1RN) exhibited over 30% of the explained variance in BMI by themselves (Fig. 2b–d), with a maximum of 37.9% variance explained (LEP). In contrast, MetBMI, ProtBMI, and ChemBMI models explained 68.9%, 70.6%, and 48.8% of the variance in BMI, respectively. Moreover, even upon eliminating several strong predictor analytes such as LEP and FABP4 from the omic datasets, the models still explained more variance in BMI than any single analyte (Supplementary Fig. 2e–h). These results indicate that the multiomic BMI prediction models explain a larger portion of the variation in BMI than any single analyte, and highlight the multivariate perturbation of blood analytes across all platforms with increasing BMI.

### Metabolic heterogeneity was responsible for the high rate of misclassification within the standard BMI classes

While the omics-inferred BMIs showed the similar phenotypic associations as the measured BMI (Fig. 1e), we observed that the difference of the predicted BMI from the measured BMI (ΔBMI) was highly correlated among the omics-based BMI models, ranging from Pearson’s *r* = 0.64 (ChemBMI vs. CombiBMI) to 0.83 (ProtBMI vs. CombiBMI) (Fig. 3a). In other words, the different omics consistently detected deviation of the omics-inferred BMI from the measured BMI per individual, implying that this deviation stemmed from a true biological signal of a perturbed physiological state rather than from noise or modeling artifacts. Actually, when individuals in the normal and obese BMI classes (defined by the WHO international standards) were subdivided by a clinical definition of metabolic health (i.e., defining metabolically unhealthy if having two or more MetS risks of the National Cholesterol Education Program (NCEP) Adult Treatment Panel III (ATP III) guidelines; see Methods)^34,35^, ΔBMI was significantly higher in MUNW and MUO groups compared to metabolically healthy, normal-weight (MHNW) and MHO groups, respectively, for all omics categories (Fig. 3b), suggesting that the deviations of model predictions are related to metabolic health.

**Figure 3.**
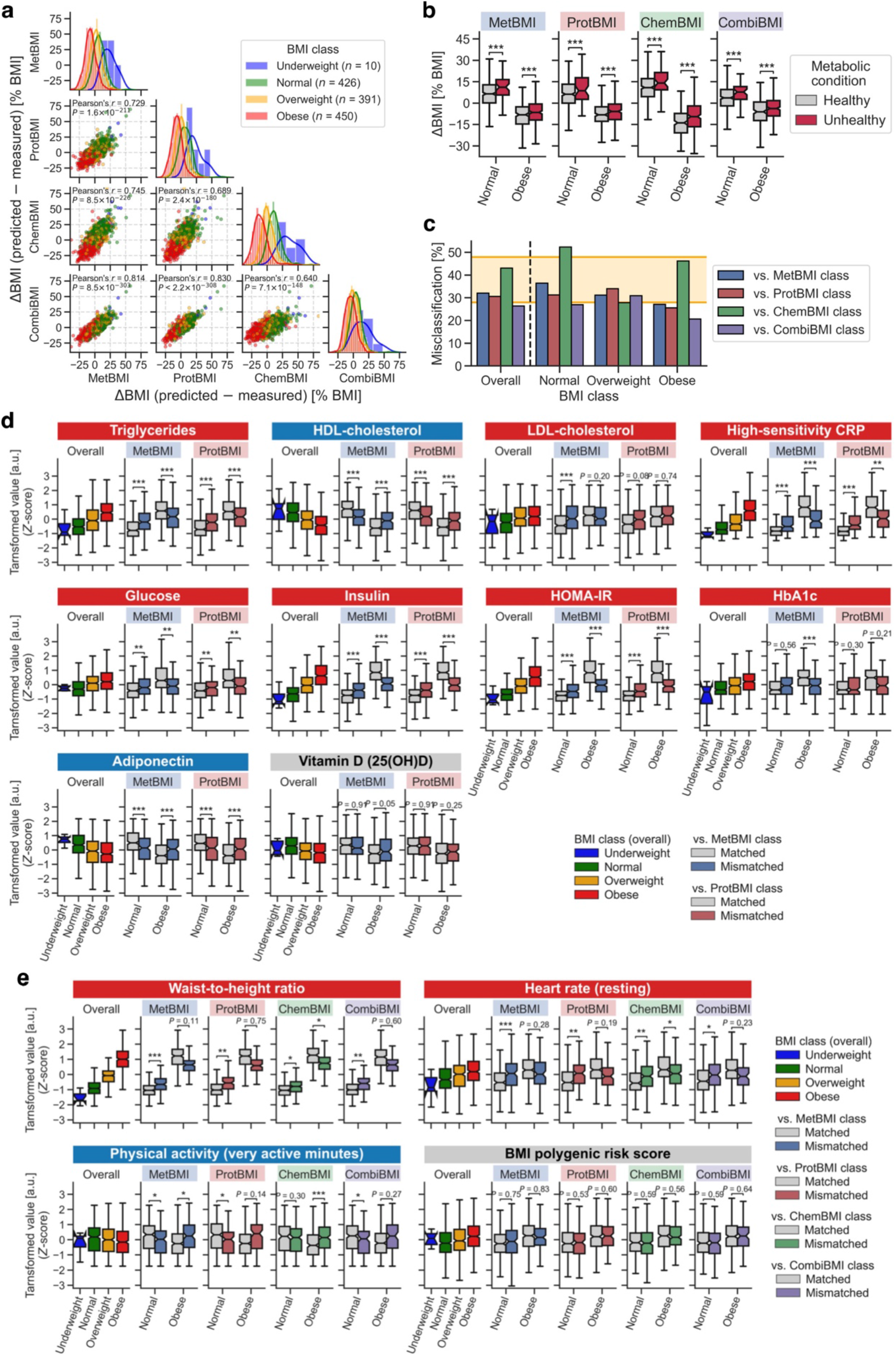
Metabolic heterogeneity was responsible for the high rate of misclassification within the standard BMI classes. **a** Difference of the omics-inferred Body Mass Index (BMI) from the measured BMI (ΔBMI). MetBMI: metabolomics-inferred BMI, ProtBMI: proteomics-inferred BMI, ChemBMI: clinical chemistries-inferred BMI, CombiBMI: combined omics-inferred BMI, *P*: adjusted *P*-value of two-sided Pearson’s correlation test with the Benjamini–Hochberg method across the six combinations, *n*: the number of participants in each BMI class (total *n* = 1,277 participants). The line in histogram panel indicates the kernel density estimate. **b** Difference in ΔBMI between clinically-defined metabolic health conditions within the normal or obese BMI class. Significance was assessed using ordinary least squares (OLS) linear regression with BMI, sex, age, and ancestry principal components (PCs) as covariates, while adjusting multiple testing with the Benjamini–Hochberg method across the eight (two BMI classes × four omics categories) regressions. **c** Misclassification rate of overall cohort or each BMI class against the omics-inferred BMI class. Range of the previously reported misclassification rate^36,37^ is highlighted with orange background. Note that the underweight BMI class is not presented due to small sample size, but its misclassification rate was 80% against CombiBMI class and 100% against the others. **d, e** Difference in the obesity-related clinical blood marker (**d**) or BMI-associated physiological feature (**e**) between Matched and Mismatched groups within the normal or obese BMI class. Significance was assessed using OLS linear regression with BMI, sex, age, and ancestry PCs as covariates, while adjusting multiple testing with the Benjamini–Hochberg method across the 40 (**d**, 2 BMI classes × 2 omics categories × 10 markers) or 216 (**e**, 2 BMI classes × 4 omics categories × 27 features) regressions. Four of the 27 features that were significantly associated with BMI (Fig. 1c) are representatively presented in **e**, and the other results are found in Supplementary Data 6. HDL: high-density lipoprotein, LDL: low-density lipoprotein, CRP: C-reactive protein, HOMA-IR: homeostatic model assessment for insulin resistance, HbA1c: glycated hemoglobin A1c, 25(OH)D: 25-hydroxyvitamin D. **b, d, e** Data: median (center line), 95% confidence interval (CI) around median (notch), [*Q*_1_, *Q*_3_] (box limits), [*x*_min_, *x*_max_] (whiskers), where *Q*_1_ and *Q*_3_ are the 1st and 3rd quartile values, and *x*_min_ and *x*_max_ are the minimum and maximum values in [*Q*_1_ − 1.5 × IQR, *Q*_3_ + 1.5 × IQR] (IQR: the interquartile range, *Q*_3_ − *Q*_1_), respectively; *n* = 373 (**b**, Healthy in Normal), 49 (**b**, Unhealthy in Normal), 208 (**b**, Healthy in Obese), 241 (**b**, Unhealthy in Obese) participants (see Supplementary Data 6 for each sample size in **d** and **e**). *Adjusted *P* < 0.05, **adjusted *P* < 0.01, ***adjusted *P* < 0.001.

Nevertheless, there has been no universally accepted definition of metabolic health^14,15,34,35^. Thus, given the high interpretability and intuitiveness of the omics-inferred BMI, we further explored a potential application: using the omics-inferred BMI (instead of the measured BMI) for improved classification of both obesity and metabolic health with the WHO international standards. Each participant was classified using each of the measured and omics-inferred BMIs based on the standard BMI cutoffs, and categorized into either Matched or Mismatched group when the measured BMI class was matched or mismatched to each omics-inferred BMI class, respectively. The misclassification rate against the omics-inferred BMI class was ∼30% across all omics categories and BMI classes (Fig. 3c), consistent with the previously reported misclassification rates about the cardiometabolic health classification^36,37^. We then examined relationships between this omics-based misclassification within normal or obese BMI class and the obesity-related clinical blood markers (Supplementary Data 6), including triglycerides, HDL-cholesterol, LDL-cholesterol, high-sensitivity C-reactive protein (hs-CRP), glucose, insulin, HOMA-IR, glycated hemoglobin A1c (HbA1c), adiponectin, and vitamin D^3,15,23,38,39^. Because ChemBMI and CombiBMI models were not independent of these markers, only the misclassification against MetBMI or ProtBMI class was examined in this analysis. The Mismatched group of normal BMI class exhibited significantly higher values of the markers that are positively associated with BMI (+_BMI_), such as triglycerides, hs-CRP, glucose, and HOMA-IR, and significantly lower values of the markers that are negatively associated with BMI (−_BMI_), such as HDL-cholesterol and adiponectin, compared to the Matched group of normal BMI class (FDR < 0.05; Fig. 3d). These patterns suggest that the participant misclassified into the normal BMI class possesses less healthy molecular profiles as similarly as the individual with overweight or obesity, corresponding to the individual with MUNW phenotype. Conversely, the Mismatched group of obese BMI class exhibited significantly lower and higher values of the positively and negatively BMI-associated markers, respectively, compared to the Matched group of obese BMI class (FDR < 0.05; Fig. 3d), suggesting that the participant misclassified as obese BMI class has healthier blood signatures, more similarly to the individual with overweight or normal-weight, corresponding to the individual with MHO phenotype. Likewise, we re-examined the 27 BMI-associated numeric physiological features (Fig. 1e, Supplementary Data 6), and found the concordant pattern of significant phenotypic differences between Matched and Mismatched groups in WHtR (+_BMI_), heart rate (+_BMI_), blood pressure (+_BMI_), and daily physical activity measures (−_BMI_) (FDR < 0.05; Fig. 3e). Importantly, there was no difference in BMI PRS (+_BMI_) between Matched and Mismatched groups (Fig. 3e), implying that lifestyle or environmental factors, rather than genetic risk, is likely involved in the discordance between the measured and omics-inferred BMIs. Furthermore, we validated and expanded these findings in the TwinsUK cohort: ΔMetBMI was significantly higher in the metabolically unhealthy group compared to the metabolically healthy group within the normal BMI class (Supplementary Fig. 6a); the misclassification rate against MetBMI class was much higher (>60%) in the normal BMI class but ∼30% in the others (Supplementary Fig. 6b); the concordant phenotypic differences between Matched and Mismatched groups were significantly observed in triglycerides (+_BMI_), HDL-cholesterol (−_BMI_), LDL-cholesterol (+_BMI_), hs-CRP (+_BMI_), and HOMA-IR (+_BMI_) (FDR < 0.05; Supplementary Fig. 6c). Remarkably, while DXA measurements were not performed in the Arivale cohort, the percentage of total fat in whole body (+_BMI_) and the ratio of fat in android region to fat in gynoid region (+_BMI_) were significantly higher in Mismatched group compared to Matched group within the normal BMI class of the TwinsUK cohort (FDR < 0.05; Supplementary Fig. 6c). Taken together, these results suggest that the omics-based BMI models can identify heterogeneous metabolic health states which are not captured by the measured BMI with the standard BMI cutoffs.

### Metabolomics-inferred BMI reflected gut microbiome profiles better than BMI

The gut microbiome has been shown to causally affect host obesity phenotypes in a mouse model^40^, and humans with obesity generally exhibit lower bacterial α-diversity (i.e., the species richness and/or evenness of an ecological community)^41,42^. However, certain meta-analyses of human case-control studies suggest an inconsistent relationship between the gut microbiome and obesity^43,44^. Given our previous finding that the association between blood metabolites and bacterial diversity is dependent on BMI^20^ and the current finding that the omics-based BMI models capture heterogeneous metabolic health states (Fig. 3), we hypothesized that MetBMI represents gut microbiome α-diversity better than the measured BMI. For the 702 Arivale participants who had both stool-derived gut microbiome and blood omic datasets (Fig. 4a; see Methods), we examined relationships between gut microbiome α-diversity (the number of observed species, Shannon’s index, and Chao1 index) and the omics-based BMI misclassification. Matched and Mismatched groups against MetBMI class showed significant differences in all α-diversity metrics within both normal and obese BMI classes (Fig. 4b), with the concordant pattern to the clinical markers and BMI-associated features (−_BMI_; e.g., HDL-cholesterol; Fig. 3d, e), implying that the MetBMI class reflects bacterial diversity better than BMI class. Interestingly, the misclassification against the other omics categories did not show these significant differences for all α-diversity metrics and both BMI classes (Fig. 4b), consistent with our previous observation that plasma metabolomics showed a much stronger correspondence to gut microbiome structure than either proteomics or clinical labs^20^.

**Figure 4.**
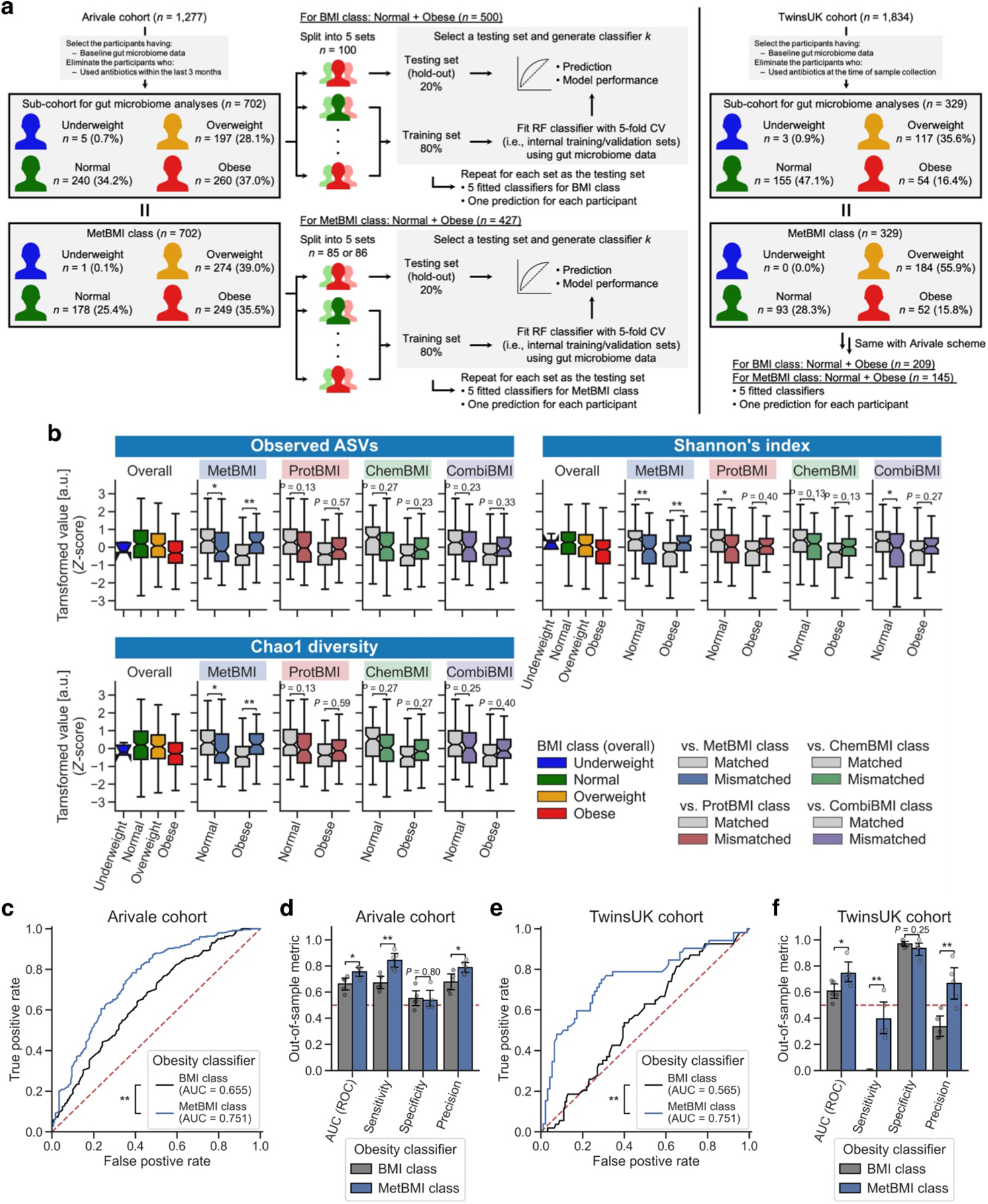
Metabolomics-inferred BMI reflected gut microbiome profiles better than BMI. **a** Overview of study cohorts and the gut microbiome-based obesity classifier generation. BMI: Body Mass Index, MetBMI: metabolomics-inferred BMI, RF: random forest, CV: cross-validation. **b** Difference in gut microbiome α-diversity between Matched and Mismatched groups within the normal or obese BMI class. Significance was assessed using ordinary least squares (OLS) linear regression with BMI, sex, age, and ancestry principal components (PCs) as covariates, while adjusting multiple testing with the Benjamini–Hochberg method across the 24 (2 BMI classes × 4 omics categories × 3 metrics) regressions. ProtBMI: proteomics-inferred BMI, ChemBMI: clinical chemistries-inferred BMI, CombiBMI: combined omics-inferred BMI, ASV: amplicon sequence variant. Data: median (center line), 95% confidence interval (CI) around median (notch), [*Q*_1_, *Q*_3_] (box limits), [*x*_min_, *x*_max_] (whiskers), where *Q*_1_ and *Q*_3_ are the 1st and 3rd quartile values, and *x*_min_ and *x*_max_ are the minimum and maximum values in [*Q*_1_ − 1.5 × IQR, *Q*_3_ + 1.5 × IQR] (IQR: the interquartile range, *Q*_3_ − *Q*_1_), respectively. *n* = 240 (Normal), 260 (Obese) participants (see Supplementary Data 6 for each sample size). *Adjusted *P* < 0.05, **adjusted *P* < 0.01. **c, e** Receiver operator characteristic (ROC) curve of the gut microbiome-based model classifying participants to the normal vs. obese class in the Arivale (**c**) or TwinsUK (**e**) cohort. Each ROC curve was generated from the overall participants: *n* = 500 (**c**, BMI class), 427 (**c**, MetBMI class), 209 (**e**, BMI class), 145 (**e**, MetBMI class) participants. The red dashed line indicates a random classification line. AUC: area under curve. \*\**P* < 0.01 in two-sided unpaired DeLong’s test. **d, f** Comparison of model performance between the BMI and MetBMI classifiers in the Arivale (**d**) or TwinsUK (**f**) cohort. Out-of-sample metric value was calculated from each corresponding hold-out testing set. Data: mean with 95% CI, *n* = 5 models. \**P* < 0.05, \*\**P* < 0.01 in two-sided Welch’s *t*-test.

We further examined the predictive power of gut microbiome profiles for MetBMI. For each of the measured BMI and MetBMI classes, we generated models classifying individuals into normal class versus obese class based on gut microbiome 16S rRNA gene amplicon sequencing data, using a fivefold iteration scheme of the RF algorithm with fivefold CV (Fig. 4a; see Methods). Compared to the classifier for the measured BMI class, the classifier for MetBMI class showed significantly larger area under curve (AUC) in the receiver operator characteristic (ROC) curve in the Arivale cohort (AUC = 0.66 (BMI), 0.75 (MetBMI); Fig. 4c), with significantly higher sensitivity and precision (Fig. 4d). Moreover, by applying the same scheme to the stool-derived whole metagenomic shotgun sequencing (WMGS) data of the 329 TwinsUK participants^45^ (Fig. 4a; see Methods), we validated that the gut microbiome-based obesity classifier for MetBMI class significantly outperformed the classifier for the measured BMI class in the TwinsUK cohort (AUC = 0.57 (BMI), 0.75 (MetBMI); Fig. 4e, f). Note that these classifiers were regenerated for the TwinsUK cohort (instead of using the classifiers that were fitted to the Arivale dataset; Fig. 4a) due to the difference in sequencing methods (amplicon sequencing vs. WMGS), while considering that the TwinsUK participants’ MetBMIs were predicted from the Arivale-fitted MetBMI models (Fig. 1a). Altogether, these findings suggest that, although other factors (e.g., dietary intake^19^) may be involved, MetBMI has a stronger correspondence to gut microbiome features than the standard BMI.

### Metabolic health of the metabolically obese group was substantially improved following a healthy lifestyle intervention

In the Arivale program, healthy lifestyle coaching was provided to all participants, resulting in clinical improvement across multiple measures of health^25^. This coaching intervention was personalized for each participant to improve the participant’s health based on the combination of clinical laboratory tests, genetic predispositions, and published scientific evidence, and administered via telephone by registered dietitians, certified nutritionists, or registered nurses (see Methods and a previous report^25^). To investigate the longitudinal changes in omic profiles during the program, we defined a sub-cohort of 608 participants based on the available longitudinal measurements (Fig. 5a; see Methods). Given the participant-dependent variability in both count and time point of data collections, we estimated the average trajectory of each measured or omics-inferred BMI in the Arivale sub-cohort using a linear mixed model (LMM) with random effects for each participant (see Methods). Consistent with the previous analysis^25,46^, the mean BMI estimate for the overall cohort decreased during the program (Fig. 5b). The decrease of MetBMI was larger than that of measured BMI, while the decrease of ProtBMI was minimal and even smaller than that of measured BMI (Fig. 5b), suggesting that plasma metabolomics is highly responsive to the lifestyle intervention in the short term, while proteomics (measured from the same blood draw) is more resistant to change during the same intervention period. Subsequently, we generated LMMs with the baseline BMI class stratification, and confirmed that a significant decrease in the mean BMI estimate was observed in the overweight and obese BMI classes, but not in the normal BMI class (Fig. 5c). Concordantly, the mean estimates of ProtBMI and ChemBMI exhibited negative changes over time in the overweight and obese BMI classes, but not in the normal BMI class (Fig. 5c). In contrast, the mean estimate of MetBMI exhibited a significant decrease across all BMI classes (Fig. 5c), suggesting that metabolomics data captures information about the metabolic health response to the lifestyle intervention, beyond the baseline BMI class and the changes in BMI and other omic profiles.

**Figure 5.**
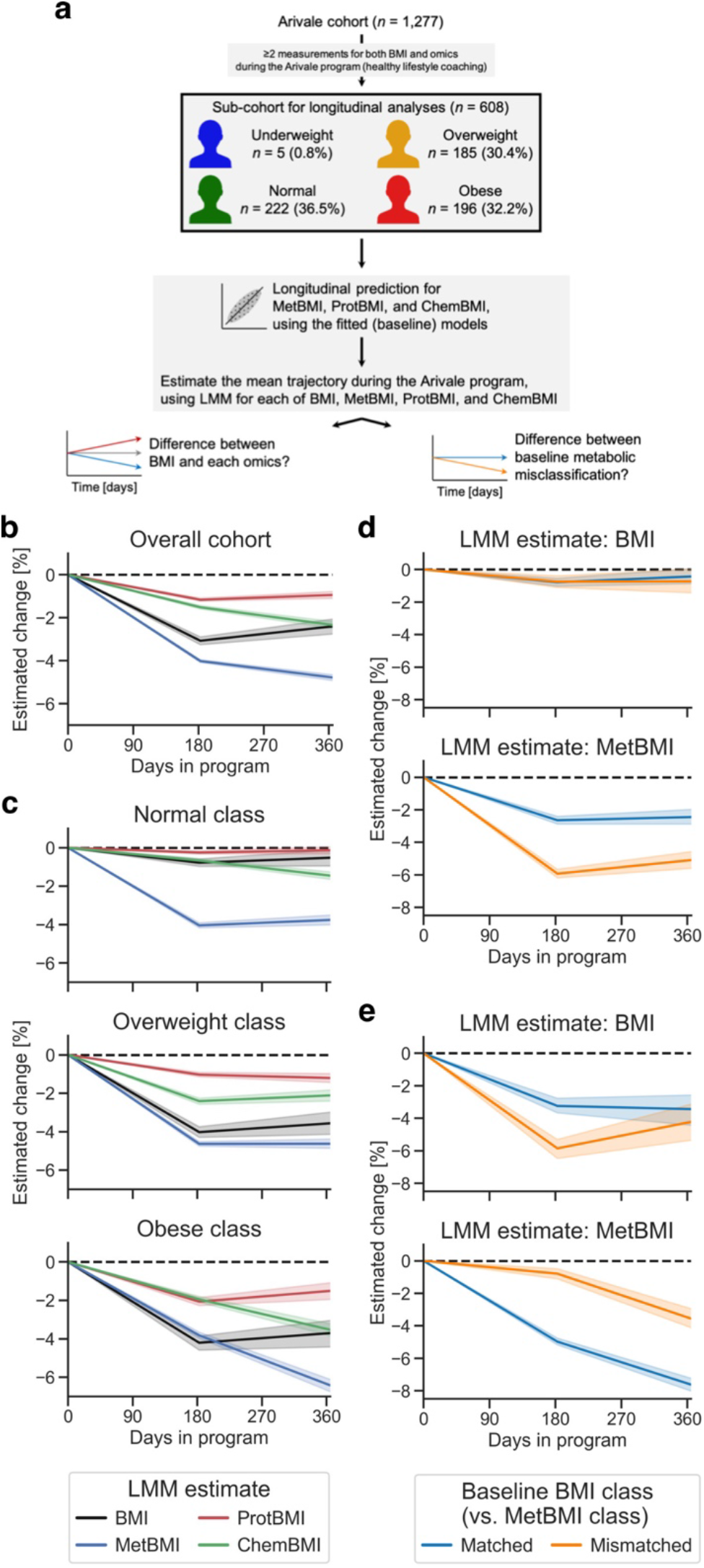
Metabolic health of the metabolically obese group was substantially improved following a healthy lifestyle intervention. **a** Overview of the longitudinal analysis using omics-inferred Body Mass Index (BMI). BMI: measured BMI, MetBMI: metabolomics-inferred BMI, ProtBMI: proteomics-inferred BMI, ChemBMI: clinical chemistries-inferred BMI, LMM: linear mixed model. **b, c** Longitudinal change in the omics-inferred BMI within the overall cohort (**b**) or within each baseline BMI class (**c**). Average trajectory of each measured or omics-inferred BMI was independently estimated using LMM with random effects for each participant (see Methods) in the overall cohort (**b**) or in each baseline BMI class-stratified group (**c**). **d, e** Longitudinal change in MetBMI of the misclassified participants within the normal (**d**) or obese (**e**) BMI class. Average trajectory of each BMI or MetBMI was independently estimated using the above LMM with the baseline misclassification of BMI class against MetBMI class as additional fixed effects (see Methods) in each baseline BMI class-stratified group. **b**–**e** The dashed line corresponds to the baseline value of each estimate. Data: mean with 95% confidence interval (CI); *n* = 608 (**b**), 222 (**c**, Normal), 185 (**c**, Overweight), 196 (**c**, Obese), 137 (**d**, Matched), 85 (**d**, Mismatched), 139 (**e**, Matched), 57 (**e**, Mismatched) participants.

Given the existence of multiple metabolic health sub-states within the standard BMI classes (Fig. 3), we further investigated the difference between misclassification strata against the baseline MetBMI class. In the (baseline) normal BMI class, while the mean estimate of the measured BMI remained constant in both Matched and Mismatched groups, the mean MetBMI estimate exhibited larger reduction in Mismatched group than Matched group (Fig. 5d), suggesting that the participants with MUNW phenotype improved their metabolic health to a greater extent than the participants with MHNW phenotype. Likewise, in the (baseline) obese BMI class, while the decrease in the mean estimate of the measured BMI was not significantly different between Matched and Mismatched groups at one year after the enrollment, the decrease in the mean MetBMI estimate was larger in Matched group than in Mismatched group (Fig. 5e), suggesting that the participants with MUO phenotype improved their metabolic health to a greater extent than the participants with MHO phenotype. Altogether, these results suggest that metabolic health was substantially improved during the program, in accordance with an individual’s baseline metabolomic state, rather than with the individual’s baseline BMI class.

### Plasma analyte correlation network in the metabolically obese group shifted toward a structure observed in metabolically healthier state following a healthy lifestyle intervention

We explored longitudinal changes in plasma analyte correlation networks, focusing on the metabolically obese group. Based on the importance of the baseline metabolomic state (Fig. 5d, e), we first assessed relationships between each plasma analyte–analyte correlation and the baseline MetBMI within the Arivale sub-cohort (Fig. 5a; 608 participants), using their interaction term in a generalized linear model (GLM; see Methods) of each analyte–analyte pair. In this type of model, the statistical test assesses whether the relationship between any two analytes is dependent on a third variable (in this case, the baseline MetBMI). Among 608,856 pairwise relationships of plasma analytes, 100 analyte–analyte correlation pairs, comprising 82 metabolites, 33 proteins, and 16 clinical laboratory tests, were significantly modified by the baseline MetBMI (FDR < 0.05; Supplementary Data 7). Subsequently, we assessed longitudinal changes of these 100 pairs within the metabolically obese group (i.e., the baseline obese MetBMI class; 182 participants), using the interaction term (i.e., interaction with days in the program) in a generalized estimating equation (GEE; see Methods) of each analyte–analyte pair. Among the 100 pairs, 27 analyte–analyte correlation pairs were significantly modified by days in the program (FDR < 0.05; Fig. 6a, Supplementary Data 7). These 27 pairs were mainly derived from metabolites (21 metabolites, 3 proteins, 3 clinical laboratory tests). One of these time-varying pairs was homoarginine and phenyllactate (PLA). Homoarginine was recently found to be a biomarker for CVD^47^ and was a robustly retained positive predictor in MetBMI and CombiBMI models (Fig. 2a, Supplementary Fig. 5a). PLA is a gut microbiome-derived phenylalanine derivative known to have antimicrobial activity and antioxidant activity^48,49^. The positive correlation between homoarginine and PLA was observed in the metabolically obese group at baseline (Fig. 6b) and became weaker in this group during the course of the intervention (Fig. 6c), implying that metabolic dysregulation specific to the metabolically obese group was somewhat improved during the program. Collectively, these findings indicate that metabolic improvement was not limited to changes in specific blood analyte concentrations but also changes in the association structure among analytes.

**Figure 6.**
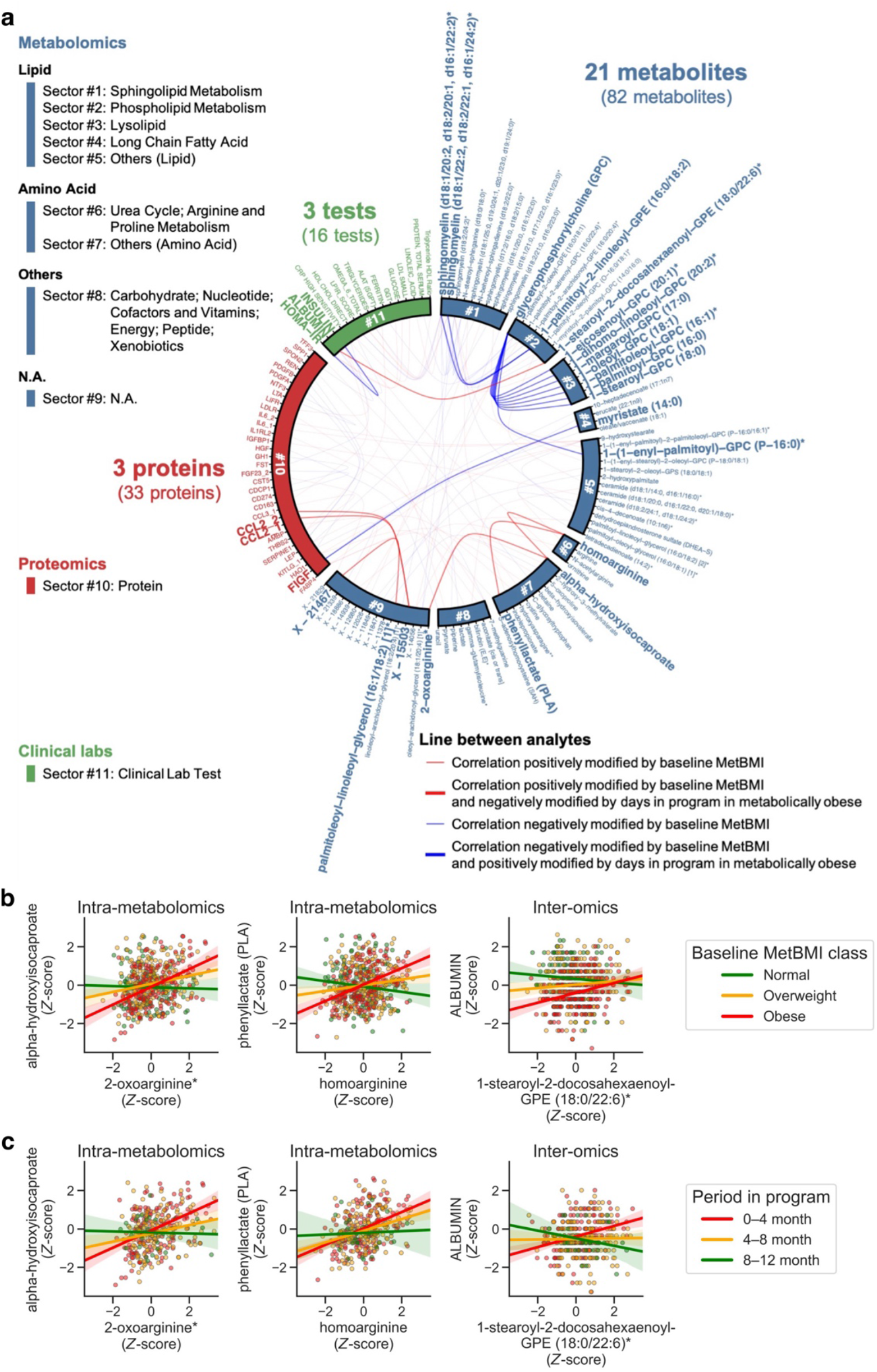
Plasma analyte correlation network in the metabolically obese group shifted toward a structure observed in metabolically healthier state following a healthy lifestyle intervention. **a** Cross-omic interactions modified by metabolomics-inferred Body Mass Index (MetBMI) and days in the program. For each of the 608,856 pairwise relationships of plasma analytes (766 metabolites, 274 proteomics, 64 clinical laboratory tests), the baseline relationship between analyte–analyte pair and MetBMI within the Arivale sub-cohort (Fig. 5a; 608 participants) was assessed using their interaction term in each generalized linear model (GLM; see Methods), while adjusting multiple testing with the Benjamini–Hochberg method. The 100 analyte–analyte pairs (82 metabolites, 33 proteins, 16 clinical laboratory tests; Supplementary Data 7) that were significantly modified by the baseline MetBMI are presented. For each of these 100 pairs, the longitudinal relationship between analyte–analyte pair and days in the program within the metabolically obese group (i.e., the baseline obese MetBMI class; 182 participants) was further assessed using their interaction term in each generalized estimating equation (GEE; see Methods), while adjusting multiple testing with the Benjamini–Hochberg method. The 27 analyte–analyte pairs (21 metabolites, 3 proteins, 3 clinical laboratory tests) that were significantly modified by days in the program are highlighted by line width and label font size. **b, c** Representative examples of the analyte–analyte pair that was significantly modified by both baseline MetBMI (**b**) and days in the program (**c**) in **a**. The solid line in each panel is the ordinary least squares (OLS) linear regression line with 95% confidence interval (CI). *n* = 530 (**b**, left), 553 (**b**, center), 566 (**b**, right) participants; *n* = 324 (**c**, left), 339 (**c**, center), 347 (**c**, right) measurements from the 182 participants of the metabolically obese group. Of note, data points outside of plot range are trimmed in these presentations.

## Discussion

Obesity is a significant risk factor for many chronic diseases^3–6^. The heterogeneous nature of human health conditions, with variable manifestation ranging from metabolic abnormalities to cardiovascular symptoms, calls for deeper molecular characterizations in order to optimize wellness and reduce the current global epidemic of chronic diseases. In this study, we have demonstrated that obesity profoundly perturbs human physiology, as reflected across all the studied omics modalities. The key findings of this study are: (1) machine learning-based multiomic BMI estimates were better suited to identifying heterogeneous metabolic health than the classically-measured BMI, while maintaining a high level of interpretability and intuitiveness attributed to the original metric (Fig. 1–3); (2) among all omics studied, metabolomic reflection of obesity exhibited the strongest correspondence to gut microbiome community structure (Fig. 4); (3) plasma metabolomics exhibited the strongest (and/or earliest) response to lifestyle coaching, while plasma proteomics exhibited a weaker (and/or more delayed) response than the measured BMI (Fig. 5b, c); (4) compared to the participants with metabolically healthy phenotype (i.e., BMI class = MetBMI class), the participants with metabolically unhealthy phenotype (i.e., BMI class < MetBMI class) exhibited a greater improvement in their metabolic health (but not in weight loss per se) in response to the healthy lifestyle coaching (Fig. 5d, e); (5) dozens of analyte–analyte associations were modified in the participants of the metabolically obese group (i.e., obese MetBMI class), following the healthy lifestyle intervention (Fig. 6).

Although BMI is used as a measure of obesity, fat distribution in the body is an important factor for understanding the heterogeneous nature of obesity. In particular, abdominal obesity, which is characterized by excessive visceral fat (rather than subcutaneous fat) around the abdominal region, is known to be associated with chronic diseases such as MetS^50^. Thus, we addressed abdominal obesity by analyzing the anthropometric WHtR^51,52^, which was highly correlated with BMI in the Arivale sub-cohort (Pearson’s *r* = 0.86; Supplementary Fig. 7a–c). We generated omics-based WHtR models (Supplementary Fig. 7a, Supplementary Data 8), and obtained consistent findings to the omics-based BMI models (Supplementary Fig. 7d–m). Interestingly, the majority of the retained analytes in each omics-based WHtR model was also retained in its corresponding omics-based BMI model with the similar feature importance (Supplementary Fig. 8a–d). In addition, ΔWHtR was highly correlated with ΔBMI across all omics categories (Supplementary Fig. 8e). Moreover, although the WC measurements were not available for the defined TwinsUK cohort, direct fat measurements of the android region by DXA were associated with MetBMI class in the TwinsUK cohort (Supplementary Fig. 6c). Therefore, although BMI requires complementary information of the WC-related measurements for the diagnosis of abdominal obesity, the omics-based BMI model likely captures the obesity characteristics including abdominal obesity.

Multiple observational studies have explored obesity biomarkers. The involvements of insulin/insulin-like growth factor (IGF) axis and chronic low-grade inflammation have been discussed in the context of obesity-related disease risks^5,6^, backed up by robust associations of obesity with IGFBP1/2 (−_BMI_), adipokines such as LEP (+_BMI_), adiponectin (−_BMI_), FABP4 (+_BMI_), and ADM (+_BMI_), and proinflammatory cytokines such as interleukin 6 (IL6; + _BMI_)^23,53^. Consistent with these well-known associations, we observed positive BMI associations with LEP, FABP4, IL1RN, IL6, ADM, and insulin and negative BMI associations with IGFBP1/2 and adiponectin (Fig. 2c, d). Importantly, all these known biomarkers were incorporated into our omics-based BMI models, and most of them were consistently retained as important features of these models (Fig. 2a; Supplementary Fig. 5b, c). At the same time, we observed that RAGE explained a relatively small proportion of the variance in BMI (Fig. 2c), while being a strong negative predictive feature in all ten models of ProtBMI and CombiBMI (Fig. 2a, Supplementary Fig. 5b). Soluble RAGE (sRAGE) has been gradually highlighted in the contexts of T2DM and CVD^54^, with several reports on the negative association between sRAGE and BMI^55^. Therefore, omics-inferred BMI may reflect not only obesity status but also the early transition towards clinical manifestations of obesity-related chronic diseases.

Likewise, many epidemiological studies have revealed metabolomic biomarkers for obesity^56,57^. In line with these previous findings, we have confirmed positive BMI associations with mannose, uric acid (urate), and glutamate and negative BMI associations with asparagine and glycine (Fig. 2b). Furthermore, all of these metabolites were consistently incorporated into all ten models of MetBMI and CombiBMI (Fig. 2a, Supplementary Fig. 5a). In addition, many lipids emerged as strong predictors in MetBMI and CombiBMI models; in particular, glycerophosphocholines (GPCs) were negative predictors in these models, while sphingomyelins (SMs) were positive predictors (Fig. 2a, Supplementary Fig. 5a), even though both have a phosphocholine group in common. Although lipid has traditionally been regarded as a factor that is positively associated with obesity, recent metabolomics studies have revealed variable trends for different fatty acid species; e.g., plasma lysophosphatidylcholines (LPCs) are decreased in mice with obesity (high-fat diet model)^58^, which corresponded well with our results (e.g., LPC(18:1), described as 1-oleoyl-GPC(18:1) in Fig. 2b and Supplementary Fig. 5b). However, because there are many combinations of acyl residues in lipids and many potential confounding factors with obesity, systematic understanding of the species-level lipid biomarkers for obesity remains challenging^56,57^. Our approach, applying machine learning to metabolomics data, addresses this challenge by automatically and systematically providing a molecular signature of obesity, reflecting the versatile and complex metabolite species. Altogether, omics-based BMI models can be regarded as multidimensional profiles of obesity, possessing detailed mechanistic information.

Recently, Cirulli and colleagues have reported a machine learning model for estimating BMI from blood metabolomics, which captured obesity-related phenotypes^21^. Their main model explained 39.1% of the variance in BMI, while our MetBMI model explained 68.9% of the variance in BMI (Fig. 2b). Other than the difference in cohorts, the performance gap is likely a result of differences in modeling strategies. Cirulli and colleagues stringently selected 49 metabolites, out of their metabolomics panel of 1,007 metabolites, based on a pre-screening for significant adjusted-associations with BMI, and subsequently applied a tenfold CV implementation of ridge or LASSO method. In contrast, we used LASSO method for feature selection, applying it to our full metabolomics panel of 766 metabolites. In addition to the increased number of metabolites included in the model fitting, our higher performance may stem from the presence of metabolites which were critical for BMI prediction in a multivariate model, but not strongly associated with BMI on their own. Actually, similarly to the above example of RAGE in ProtBMI model, our MetBMI model contained multiple metabolites that were weakly associated with BMI but consistently retained across all ten models (Fig. 2b, Supplementary Fig. 5a). At the same time, the majority of the 49 metabolites reported by Cirulli and colleagues (14–20 metabolites among the 31–41 corresponding metabolites in our metabolomics panel) were retained in at least one of the ten MetBMI models. Therefore, our strategy of feature selection through machine learning, without a pre-filtering step, may be preferable for predicting BMI from metabolomics.

A recent study investigating multiomic changes in response to weight perturbations demonstrated that some weight gain-associated blood signatures were reversed during subsequent weight loss, while others persisted^59^. Interestingly, we found that MetBMI was more responsive to the healthy lifestyle intervention than the measured BMI or ChemBMI, while ProtBMI was more resistant to the same intervention (Fig. 5b, c). Our analyses of the predictors in the omics-based BMI models (Fig. 2; Supplementary Fig. 2e–h, 5) suggested that the distribution of feature importance among metabolites was considerably wider, while only a small subset of measured proteins (∼5 proteins) was predominantly reflective of obesity profiles. Therefore, the effect of lifestyle coaching may consist of small additive contributions in blood metabolites in the short term. However, a longer longitudinal analysis is needed to infer the physiological meaning of these omics-dependent dynamics. For instance, it is possible that ProtBMI shows a delayed response to weight loss (over a span greater than a year measured presently; Fig. 5b, c), indicating blood metabolites and proteins may be early and late responders to a lifestyle intervention, respectively, such as in the case of the changes in blood glucose compared to the changes in HbA1c when assessing glucose homeostasis^60^. If the difference between the measured and omics-inferred BMIs remains constant even after one year, we would conclude that blood metabolites and proteins are more and less sensitive to weight loss than the measured BMI, respectively. In either scenario, monitoring blood multiomics during weight loss programs could help participants maintain their motivation to stay engaged with persistent lifestyle changes, because they would receive rapid feedback on how lifestyle changes were impacting their health, even in the absence of weight loss. In addition, long-term maintenance of the improvement is an important challenge for lifestyle interventions; although there is variability between prior reports, one study estimated that only ∼20% of the individuals with overweight successfully maintain their weight loss in post-intervention^61^. Despite this relatively low rate of long-term success, there is evidence that lifestyle interventions had benefits in preventing diabetes incidence as far as 20 years post-intervention, even if weight was regained^62,63^. The observed larger improvement of MetBMI compared to the measured BMI could potentially contribute to this protective long-term effect, persisting even when weight is regained. Further investigation is required, especially with regard to the long-term dynamics of MetBMI and ProtBMI responses, which may provide a foothold in developing scientific strategies aimed at long-term maintenance of metabolic health.

Despite a number of highly promising findings, there were several limitations to our study. For example, this study was not designed as a randomized control trial, and we cannot strictly evaluate the effectiveness of the lifestyle intervention (e.g., bigger improvements in the obese group compared to the normal-weight group may be due to the regression-toward-the-mean effect^46^). In addition, we used time as the variable in longitudinal analyses under an assumption that the program enrollment itself affected participant’s BMI and omic profiles. However, if we had more detailed data on the intervention (e.g., magnitude, participants’ compliance), we would be able to improve the assessment of its effect. The generalizability of our findings may be limited, because this study was an observational study of largely Caucasian cohorts from the Pacific West of the U.S. and from the U.K. and because validation with an external cohort relied on the female-dominated cohort (96.7%) and its metabolomics data. Our measurements did not cover all biomolecules in blood; in particular, proteomics was based on three targeted Olink panels. Thus, our findings on metabolomic and proteomic states are restricted to the analytes that we could measure. Nevertheless, this study will serve as a valuable resource for robustly characterizing metabolic health from the blood and identifying actionable targets for health management.

## Methods

### Study cohort

The main study cohort (Arivale cohort) was derived from 6,223 individuals who participated in a wellness program offered by a currently closed commercial company (Arivale Inc., Washington, USA) between 2015–2019. An individual was eligible for enrollment if the individual was over 18 years old, not pregnant, and a resident of any U.S. state except New York; participants were primarily recruited from Washington, California, and Oregon. The participants were not screened for any particular disease. During the Arivale program, each participant was provided personalized lifestyle coaching via telephone by registered dietitians, certified nutritionists, or registered nurses. This coaching was designed to improve the participant’s health based on the combination of clinical laboratory tests, genetic predispositions, and published scientific evidence; e.g., reduction of sodium intake might be recommended to any participants with high blood pressure, but if they also had risk alleles indicating enhanced susceptibility to dietary sodium, this risk would be emphasized (see a previous report^25^ for more details). In the current study, to compare the associations between Body Mass Index (BMI) and host phenotypes across different omics, we limited the original cohort to the participants whose datasets contained (1) all main omic measurements (metabolomics, proteomics, clinical laboratory tests) from the same first blood draw, (2) a BMI measurement within ±1.5 month from the first blood draw, and (3) genetic information (for using as covariates). We also eliminated (1) outlier participants whose baseline BMI was beyond ±3 s.d. from the mean in the baseline BMI distribution and (2) participants whose any of omic datasets contained more than 10% missingness in the filtered analytes (see the next section). The final Arivale cohort consisted of 1,277 (821 female and 456 male) participants (Fig. 1a), which exhibited consistent demographics (Supplementary Fig. 1a–c, Supplementary Data 1) with the study cohorts defined in the previous Arivale studies^20,25–29^. For the analyses of gut microbiome, sub-cohort was defined with the 702 (486 female and 216 male) participants from the Arivale cohort, who collected a stool sample within ±1.5 month from the first blood draw and did not use antibiotics in the last three months (Fig. 4a, Supplementary Data 1). For longitudinal analyses, sub-cohort was defined with the 608 (410 female and 198 male) participants from the Arivale cohort, whose datasets contained two or more time-series datasets for both BMI and omics during 18 months after enrollment (Fig. 5a, Supplementary Data 1). For the analyses of waist-to-height ratio (WHtR), sub-cohort was defined with the 1,078 (689 female and 389 male) participants from the Arivale cohort, whose datasets contained the baseline WHtR measurement within ±1.5 month from the first blood draw and within ±3 s.d. from the mean in the baseline WHtR distribution (Supplementary Fig. 7a, Supplementary Data 1).

The external cohort (TwinsUK cohort) was derived from 17,630 individuals who participated in the TwinsUK Registry, a British national register of adult twins^31^. Twins were recruited as volunteers by media campaigns without screening for any particular disease. The participants had two or more clinical visits for biological sampling between 1992–2022. In the current study, to validate our findings in the Arivale cohort, we limited the original cohort to the participants whose datasets contained all measurements for metabolomics^32^, BMI, and the obesity-related standard clinical measures (i.e., defined by triglycerides, high-density lipoprotein (HDL)-cholesterol, low-density lipoprotein (LDL)-cholesterol, glucose, insulin, and homeostatic model assessment for insulin resistance (HOMA-IR) throughout the current study) from the same visit. We also eliminated (1) outlier participants whose BMI was beyond ±3 s.d. from the mean in the overall BMI distribution and participants whose metabolomic dataset contained more than 10% missingness in the filtered metabolites (see the next section). The final TwinsUK cohort consisted of 1,834 (1,774 female and 60 male) participants (Fig. 1a, Supplementary Fig. 1d–f, Supplementary Data 1). For the analyses of gut microbiome, sub-cohort was defined with the 329 (307 female and 22 male) participants from the TwinsUK cohort, who collected a stool sample within ±1.5 month from the clinical visit and did not use antibiotics at that time (Fig. 4a, Supplementary Data 1).

The current study was conducted with de-identified data of the participants who had consented to the use of their anonymized data in research. All procedures were approved by the Western Institutional Review Board (WIRB) with Institutional Review Board (IRB) (Study Number: 20170658 at Institute for Systems Biology and 1178906 at Arivale) and by the TwinsUK Resource Executive Committee (TREC) (Project Number: E1192).

### Data collections and data cleaning

Multiomics data for the Arivale participants included genomics and longitudinal measurements of metabolomics, proteomics, clinical laboratory tests, gut microbiomes, wearable devices, and health/lifestyle questionnaires. Peripheral venous blood draws for all measurements were performed by trained phlebotomists at LabCorp (Laboratory Corporation of America Holdings, North Carolina, USA) or Quest (Quest Diagnostics, New Jersey, USA) service centers. Saliva to measure analytes such as diurnal cortisol and dehydroepiandrosterone (DHEA) was sampled by participants at home using a standardized kit (ZRT Laboratory, Oregon, USA). Likewise, stool samples for gut microbiome measurements were obtained by participants at home using a standardized kit (DNA Genotek, Inc., Ottawa, Canada).

#### Genomics

DNA was extracted from each whole blood sample and underwent whole genome sequencing (1,257 participants) or single-nucleotide polymorphisms (SNP) microarray genotyping (20 participants). Genetic ancestry was calculated with principal components (PCs) using a set of ∼100,000 ancestry-informative SNP markers, as described previously^25^. Polygenic risk scores (PRSs) were constructed using publicly available summary statistics from published genome-wide association studies (GWAS), as described previously^27^.

#### Blood-measured omics

Metabolomics data was generated by Metabolon, Inc. (North Carolina, USA), using ultra-high-performance liquid chromatography-tandem mass spectrometry (UHPLC-MS/MS) for plasma derived from each whole blood sample. Proteomics data was generated using proximity extension assay (PEA) for plasma derived from each whole blood sample with several Olink Target panels (Olink Proteomics, Uppsala, Sweden), and only the measurements with the Cardiovascular II, Cardiovascular III, and Inflammation panels were used in the current study since the other panels were not necessarily applied to all samples. All clinical laboratory tests were performed by LabCorp or Quest in a Clinical Laboratory Improvement Amendments (CLIA)-certified lab, and only the measurements by LabCorp were selected in the current study to eliminate potential differences between vendors. In the current study, the batch-corrected datasets with in-house pipeline were used, and metabolomic dataset was log_*e*_-transformed. In addition, analytes missing in more than 10% of the baseline samples were removed from each omic dataset, and observations missing in more than 10% of the remaining analytes were further removed. The final filtered metabolomics, proteomics, and clinical labs consisted of 766 metabolites, 274 proteins, 71 clinical laboratory tests, respectively (Supplementary Data 2).

#### Gut microbiome

Gut microbiome data was generated based on 16S amplicon sequencing of the V3+V4 region using a MiSeq sequencer (Illumina, Inc., California, USA) for DNA extracted from each stool sample, as previously described^28^. Briefly, the FASTQ files were processed using the mbtools workflow (https://github.com/Gibbons-Lab/mbtools) to remove noise, infer amplicon sequence variants (ASVs), and remove chimeras. Taxonomy assignment was performed using the SILVA ribosomal RNA gene database (version 132)^64^. In the current study, the final collapsed ASV table across the samples consisted of 394, 341, 85, 45, 26, and 16 taxa for species, genus, family, order, class, and phylum, respectively. Gut microbiome α-diversity was calculated at the ASV level using Shannon’s index calculated by 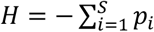,where *p*_*i*_ is the proportion of a community *i* represented by ASVs, or using Chao1 diversity score calculated by 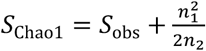, where *S*_*obs*_ is the number of observed ASVs, *n*_*1*_ is the number of singletons (ASVs captured once), and *n*_*2*_ is the number of doubletons (ASVs captured twice).

#### Anthropometrics, saliva-measured analytes, and daily physical activity measures

Anthropometrics including weight, height, and waist circumference (WC) and blood pressure were measured at the time of blood draw and also reported by participants, which generated diverse timing and number of observations depending on each participant. BMI and WHtR were simultaneously calculated from the measured anthropometrics with the weight divided by squared height [kg m^−2^] and the WC divided by height [unitless], respectively. Measurements of saliva samples were performed in the testing laboratory of ZRT Laboratory. Daily physical activity measures such as heart rate, moving distance, step count, burned calories, floors climbed, and sleep quality were tracked using the Fitbit wearable device (Fitbit, Inc., California, USA). To manage variations between days, monthly averaged data was used for these daily measures. In the current study, the baseline measurement for these longitudinal measures was defined with the closest observation to the first blood draw per participant and data type, and each dataset was eliminated from analyses when its baseline measurement was beyond ±1.5 month from the first blood draw.

Data resource for the TwinsUK participants included longitudinal measurements of metabolomics, clinical laboratory tests, dual-energy X-ray absorptiometry (DXA), and health/lifestyle questionnaires^31^. The necessary datasets for the current study were provided by Department of Twin Research & Genetic Epidemiology (King’s College London). In the current study, after each provided dataset was cleaned as follows, the earliest visit among the visits from which all of metabolomics, BMI, and the standard clinical measures had been measured was defined as the baseline visit for each participant. As exception, the later visit among them was prioritized as the baseline visit, if the participant had gut microbiome data within ±1.5 month from the visit. Only the baseline visit measurements were analyzed.

#### Blood-measured metabolomics

Metabolomics data was originally generated by Metabolon, Inc., using UHPLC-MS/MS for each serum sample^32^. In the current study, the provided median-normalized dataset was log_*e*_-transformed. In addition, metabolites missing in more than 10% of the overall samples were removed from metabolomic dataset, and observations missing in more than 10% of the remaining metabolites were further removed. The final filtered metabolomics consisted of 683 metabolites.

#### BMI

In the current study, the BMI values that had been already calculated and included in the provided metabolomics data file were used.

#### Standard clinical measures and other phenotypic measures

In the current study, because the provided phenotypic datasets contained multiple measurements for a phenotype even from a single visit of a participant (e.g., due to project difference, repeated measurements), multiple measurements were flattened into a single measurement for a phenotype per each participant’s visit by taking the mean value. During this flattening step, difference in unit was properly adjusted, and the value indicating below detection limit was regarded as zero. HOMA-IR was calculated from the datasets of glucose, insulin, and fasting condition with the formula: HOMA-IR = fasting glucose [mmol L^−1^] × fasting insulin [mIU L^−1^] × 22.5^−1^.

#### Gut microbiome

Gut microbiome data was originally generated based on whole metagenomic shotgun sequencing (WMGS) using a HiSeq 2500 sequencer (Illumina, Inc.) for DNA extracted from each stool sample^45^. In the current study, the raw sequencing data was obtained from the National Center for Biotechnology Information (NCBI) Sequence Read Archive (SRA) project (PRJEB32731), and applied to a processing pipeline (https://github.com/Gibbons-Lab/pipelines). Briefly, the obtained FASTQ files were processed using the fastp (version 0.23.2) tool^65^ to filter and trim the reads, and taxonomic abundance was obtained using the Kraken 2 (version 2.1.2) and Bracken (version 2.6.0) tools^66^ with the Kraken 2 default database (based on NCBI RefSeq). The final collapsed taxonomic table across the samples consisted of 4,669, 1,225, 354, 167, 76, and 35 taxa for species, genus, family, order, class, and phylum, respectively.

### Blood omics-based BMI and WHtR models

For each Arivale baseline omic dataset, missing values were first imputed with a random forest (RF) algorithm using Python missingpy (version 0.2.0) library (corresponding to R MissForrest package^67^). For sex-stratified models (Supplementary Fig. 2d), the datasets after imputation were divided into sex-stratified datasets. Subsequently, the values in each omic dataset were standardized with *Z*-score using the mean and s.d. per analyte. Then, ten iterations of least absolute shrinkage and selection operator (LASSO) modeling with tenfold cross-validation (CV) (Fig. 1a, Supplementary Fig. 7a) were performed for the (unstandardized) log_*e*_-transformed BMI or WHtR and each processed omic dataset, using *LassoCV* application programming interface (API) of Python scikit-learn (version 1.0.1) library. Training and testing (hold-out) sets were generated by splitting participants into ten sets with one set as a testing (hold-out) set and the remaining nine sets as a training set, and iterating all combinations over those ten sets; i.e., overfitting was controlled using tenfold iteration with ten testing (hold-out) sets, and hyperparameter was decided using tenfold CV with internal training and validation sets from each training set. Consequently, this procedure generated ten fitted sparse models for each omics category (Supplementary Data 3) and one single testing (hold-out) set-derived prediction from each omics category for each participant. The same modeling scheme while replacing LASSO with elastic net (EN), ridge, or RF was performed using Python scikit-learn *ElasticNetCV, RidgeCV*, or *RandomForestRegressor*-implemented *GridSearchCV* API, respectively. In this RF-modeling, the number of trees in the forest and the number of features were set as the hyperparameters to be decided through CV. For the standard measures-based models, the above modeling scheme was applied to ordinary least squares (OLS) linear regression with sex, age, triglycerides, HDL-cholesterol, LDL-cholesterol, glucose, insulin, and HOMA-IR as regressors, using Python scikit-learn *LinearRegression* API. Of note, ten split sets were fixed among the omics categories and the modeling methods, and no significant difference in BMI, WHtR, sex, age, and ancestry PC1–5 among those ten sets was confirmed, using Pearson’s *χ*^2^ test for categorical variable and Analysis of Variance (ANOVA) for numeric variable while adjusting multiple testing with the Benjamini–Hochberg method across the tested variables (Supplementary Data 1).

For the TwinsUK cohort, metabolomic dataset was applied to RF imputation and then each dataset of metabolomics and the standard clinical measures was applied to *Z*-score standardization, as well as the Arivale datasets. Utilizing the ten LASSO or OLS linear regression models that were fitted by the Arivale dataset, one single prediction was calculated from each processed dataset for each participant by taking the mean of ten predicted values. For metabolomics, ten metabolomics-based BMI (MetBMI) models were regenerated while restricting the input Arivale metabolomics to the common 489 metabolites in the Arivale and TwinsUK panels (Supplementary Fig. 3).

For the LASSO-modeling iteration analysis (Supplementary Fig. 2e–h, 7f–i), ten LASSO models were repeatedly generated with the above modeling scheme. At the end of each iteration, the variable that was retained across ten models and that had the highest absolute value for the mean of ten *β*-coefficients was removed from the input omic dataset.

For longitudinal predictions of the Arivale sub-cohort, one single prediction at a time point was calculated from each processed time-series omic dataset for each participant, utilizing the baseline LASSO model for which the participant was included in the baseline testing (hold-out) set. This was because (1) the baseline measurements were minimally affected by the personalized lifestyle coaching, (2) both count and time point of data collections were different among the participants, and (2) potential data leakage might be derived from the relationships between the baseline and following measurements for the same participant. For processing, each time-series omic dataset was applied to two-step RF imputation, where the baseline missingness was first imputed based on the baseline data structure and the remaining missingness was next imputed based on the overall data structure, and subsequently applied to *Z*-score standardization using the mean and s.d. in the baseline distribution.

Model performance was conservatively evaluated by the out-of-sample *R*^2^ that was calculated from each corresponding hold-out testing set in the Arivale cohort or from the external testing set in the TwinsUK cohort. Pearson’s *r* between the measured and predicted values was calculated from the overall participants of the Arivale or TwinsUK cohort. Difference of the predicted value from the measured value (ΔMeasure; i.e., ΔBMI or ΔWHtR) was calculated with (the predicted value – the measured value) × (the measured value)^−1^ × 100 (i.e., the unit of ΔMeasure was [% Measure]). In the RF model, the importance of a feature was calculated as the normalized total reduction of the mean squared error that was brought by the feature.

### Health classification

Each participant was classified using each of the measured and omics-inferred BMIs based on the World Health Organization (WHO) international standards for BMI cutoffs (underweight: <18.5 kg m^−2^, normal: 18.5–25 kg m^−2^, overweight: 25–30 kg m^−2^, obese: ≥30 kg m^−2^)^12^. For the misclassification of BMI class against the omics-inferred BMI class, each participant was categorized into either Matched or Mismatched group when the measured BMI class was matched or mismatched to each omics-inferred BMI class, respectively.

For a clinically-defined metabolic health classification, the participants having two or more metabolic syndrome (MetS) risks of the National Cholesterol Education Program (NCEP) Adult Treatment Panel III (ATP III) guidelines were judged as the metabolically unhealthy group, while the other participants were judged as the metabolically healthy group^34,35^. Concretely, the MetS risk components were (1) systolic blood pressure ≥130 mm Hg, diastolic blood pressure ≥85 mm Hg, or using antihypertensive medication, (2) fasting triglyceride level ≥150 mg dL^−1^, (3) fasting HDL-cholesterol level <50 mg dL^−1^ for female and <40 mg dL^−1^ for male or using lipid-lowering medication, and (4) fasting glucose level ≥100 mg dL^−1^ or using antidiabetic medication. Only the participants who had all these information were assessed in the corresponding analyses (Fig. 3b; Supplementary Fig. 6a, 7m).

### Gut microbiome-based models for classifying obesity

For the Arivale gut microbiome dataset, the whole ASV table (907 taxa from species to phylum) was preprocessed (i.e., positively shifted by one, log_*e*_-transformed, and standardized with *Z*-score using the mean and s.d. per taxon) and then applied to dimensionality reduction using *PCA* API of Python scikit-learn (version 1.0.1) library; the projected values onto the first 50 PCs (0.4–5.1% variance explained) were supplied as the input gut microbiome features. Two types of classifiers were trained on these gut microbiome features: one predicting whether an individual is obese BMI class and the other predicting whether an individual is obese MetBMI class. Both models were independently constructed through a fivefold iteration scheme of RF with fivefold CV (Fig. 4a), using Python scikit-learn *RandomForestClassifier*-implemented *GridSearchCV* API. In this RF-modeling, the number of trees in the forest and the number of features were set as the hyperparameters to be decided through CV. Training and testing (hold-out) sets were generated by splitting the participants of the normal and obese classes into five sets with one set as a testing (hold-out) set and the remaining four sets as a training set, and iterating all combinations over those five sets; i.e., overfitting was controlled using fivefold iteration with five testing (hold-out) sets, and hyperparameters were decided using fivefold CV with internal training and validation sets from each training set. Consequently, this procedure generated five fitted classifiers for each BMI or MetBMI class and one single testing (hold-out) set-derived prediction from each classifier type for each participant. Note that this prediction included two types: either normal or obese class by a vote of the trees (i.e., binary prediction) and the mean probability of obese class among the trees.

For the TwinsUK gut microbiome dataset, the whole taxonomic table (6,526 taxa from species to phylum) was preprocessed and then applied to dimensionality reduction, as well as the Arivale dataset; the projected values onto the first 50 PCs (0.2–40.1% variance explained) were supplied as the input gut microbiome features. Then, the five obesity classifiers for each BMI or MetBMI class were generated as well as the above Arivale procedure, and one single testing (hold-out) set-derived prediction from each classifier type was calculated for each participant (Fig. 4a).

Model performance of each classifier was conservatively evaluated using each corresponding hold-out testing set. Area under curve (AUC) in the receiver operator characteristic (ROC) curve and the average precision were calculated using the probability predictions, while sensitivity and specificity were calculated from confusion matrix using the binary predictions. The overall ROC curve and its AUC was calculated from all the participant’s probability predictions, using R pROC (version 1.18.0) package^68^.

### Longitudinal changes in the measured and omics-inferred BMIs

A linear mixed model (LMM) was generated for each log_*e*_-transformed measured or omics-inferred BMI in the Arivale sub-cohort, following the previous approach^25^. As fixed effects regarding time, linear regression splines with knots at 0, 6, 12, and 18 months were applied to days in program to fit time as a continuous variable rather than a categorical variable, because both count and time point of data collections were different among the participants. In addition to the linear regression splines of time as fixed effects, the LMM included sex, baseline age, ancestry PC1–5, and meteorological seasons as fixed effects (to adjust potential confounding effects) and random intercepts and random slopes of days in the program as random effects for each participant. Additionally, the same LMM for each measured or omics-inferred BMI was independently generated from each baseline BMI class-stratified group. Of note, this stratified LMM was not generated from the underweight group because its sample size was too small for convergence. For comparing difference between the misclassification strata against the baseline MetBMI class, the above LMM while adding additional fixed effects, the categorical baseline misclassification of BMI class against MetBMI class (i.e., binary for Matched vs. Mismatched) and its interaction terms with the linear regression splines of time, was generated for each measured BMI or MetBMI from each baseline BMI class-stratified group. All LMMs were modeled using *MixedLM* API of Python statsmodels (version 0.13.0) library.

### Plasma analyte correlation network analysis

Prior to the analysis, outlier values which were beyond ±3 s.d. from the mean in the Arivale sub-cohort baseline distribution were eliminated from the dataset per analyte, and seven clinical laboratory tests which became almost invariant across the participants were eliminated from analyses, allowing convergence in the following modeling. Per each analyte, values were converted with a transformation pipeline producing the lowest skewness (e.g., no transformation, the logarithm transformation for right skewed distribution, the square root transformation with mirroring for left skewed distribution) and standardized with *Z*-score using the mean and s.d.

Against 608,856 pairwise combinations of the analytes (766 metabolites, 274 proteomics, 64 clinical laboratory tests), generalized linear models (GLMs) for the baseline measurements of the Arivale sub-cohort (Fig. 5a; 608 participants) were independently generated with the Gaussian distribution and identity link function using *glm* API of Python statsmodels (version 0.13.0) library. Each GLM consisted of an analyte as dependent variable, another analyte and the baseline MetBMI as independent variables with their interaction term, and sex, baseline age, and ancestry PC1–5 as covariates. The analyte–analyte correlation pair that was significantly modified by the baseline MetBMI was obtained based on the *β*-coefficient (two-sided *t*-test) of the interaction term between the independent variables in GLM, while adjusting multiple testing with the Benjamini–Hochberg method (false discovery rate (FDR) < 0.05).

Against the significant 100 pairs from the GLM analysis (82 metabolites, 33 proteins, and 16 clinical laboratory tests; Supplementary Data 7), generalized estimating equations (GEEs) for the longitudinal measurements of the metabolically obese group (i.e., the baseline obese MetBMI class; 182 participants) were independently generated with the exchangeable covariance structure using Python statsmodels *GEE* API. Each GEE consisted of an analyte as dependent variable, another analyte and days in the program as independent variables with their interaction term, and sex, baseline age, ancestry PC1–5, and meteorological seasons as covariates. The analyte–analyte correlation pair that was significantly modified by days in the program was obtained based on the *β*-coefficient (two-sided *t*-test) of the interaction term between the independent variables in GEE, while adjusting multiple testing with the Benjamini–Hochberg method (FDR < 0.05).

### Statistical analysis

All data preprocessing and statistical analyses were performed using Python NumPy (version 1.18.1 or 1.21.3), pandas (version 1.0.3 or 1.3.4), SciPy (version 1.4.1 or 1.7.1) and statsmodels (version 0.11.1 or 0.13.0) libraries, except for using R pROC (version 1.18.0) package^68^ for DeLong’s test^69^. All statistical tests were performed using a two-sided hypothesis. In all cases of multiple testing, *P*-value was adjusted with the Benjamini–Hochberg method. Of note, because some hypotheses were not completely independent (e.g., between combined omics and each individual omics; between glucose, insulin, and HOMA-IR), this simple *P*-value adjustment was regarded as a conservative approach.

Significance was based on *P* < 0.05 for single testing and FDR < 0.05 for multiple testing. Test summaries (e.g., sample size, degrees of freedom, test statistic, exact *P*-value) are found in Supplementary Data 4, 5, 6, 9, and 10. Correlations (Fig. 1b, 3a; Supplementary Fig. 3b–d, 4b, 4f, 7c, 7d, 7l, 8d, 8e) were independently assessed using Pearson’s correlation test (Python SciPy *pearsonr* API), with the *P*-value adjustment if multiple testing. Comparisons of model performance (Fig. 1c, 1d, 4d, 4f; Supplementary Fig. 2d, 4a, 7e) were independently assessed using Welch’s *t*-test (Python statsmodels *ttest_ind* API), with the *P*-value adjustment if multiple testing. Comparison of overall ROC curves (Fig. 4c, 4e) was assessed using unpaired DeLong’s test^69^.

In all regression analyses, only the baseline datasets were used, and, unless otherwise specified, all numeric variables were centered and scaled in advance. For the Arivale datasets of anthropometrics, saliva-measured analytes, daily physical activity measures, and PRSs, (1) outlier values which were beyond ±3 s.d. from the mean in the cohort distribution were eliminated from the dataset per variable, (2) variables which became almost invariant across the participants were eliminated from the datasets, (3) values were converted with a transformation pipeline producing the lowest skewness (e.g., no transformation, the logarithm transformation for right skewed distribution, the square root transformation with mirroring for left skewed distribution), and (4) the transformed values were standardized with *Z*-score using the mean and s.d.; these preprocessed 51 variables were used as the numeric physiological features (Supplementary Data 4). Likewise, the Arivale datasets of the obesity-related clinical blood markers (i.e., selected clinical labs; Supplementary Data 6) and the TwinsUK datasets of the obesity-related phenotypic measures (Supplementary Data 6) were preprocessed. For gut microbiome α-diversity metrics, the number of observed ASVs and Chao1 index were converted with square root transformation while Shannon’s index was converted with square transformation, and then these transformed values were standardized with *Z*-score using the mean and s.d. Relationships of the numeric physiological features with the measured or omics-inferred BMI (Fig. 1e) were independently assessed using each OLS linear regression model with the (unstandardized) log_*e*_-transformed measured or omics-inferred BMI as dependent variable, a feature as independent variable, and sex, age, and ancestry PC1–5 as covariates, while adjusting multiple testing across the 255 (51 features × 5 BMI types) regressions. Relationships between Measure (i.e., BMI or WHtR) and the analytes that were retained in at least one of ten LASSO models (Fig. 2b–d, Supplementary Fig. 7k) were independently assessed using each OLS linear regression model with the (unstandardized) log_*e*_-transformed Measure as dependent variable, an analyte as independent variable, and sex, age, and ancestry PC1–5 as covariates, while adjusting multiple testing across the 210 (Fig. 2b), 75 (Fig. 2c), 42 (Fig. 2d), or 289 (Supplementary Fig. 7k) regressions. In this regression analysis, a model including the omics-inferred Measure as independent variable was also assessed as reference. Differences in ΔMeasure (i.e., ΔBMI or ΔWHtR) between clinically-defined metabolic health conditions (Fig. 3b; Supplementary Fig. 6a, 7m) were independently assessed using each OLS linear regression model with ΔMeasure as dependent variable, metabolic condition (i.e., Healthy vs. Unhealthy) as categorical independent variable, and Measure, sex, age, and ancestry PC1–5 as covariates, while adjusting multiple testing across the eight (two BMI classes × four omics categories; Fig. 3b, Supplementary Fig. 7m) or four (two BMI classes × two cohorts; Supplementary Fig. 6a) regressions. Differences in the obesity-related clinical blood markers, the BMI-associated numeric physiological features, or the gut microbiome α-diversity metrics between the misclassification strata against the omics-inferred BMI class (Fig. 3d, 3e, 4b; Supplementary Fig. 6c) were independently assessed using each OLS linear regression model with a marker, feature, or metric as dependent variable, misclassification (i.e., Matched vs. Mismatched) as categorical independent variable, and BMI, sex, age, and ancestry PC1–5 as covariates, while adjusting multiple testing across the 40 (2 BMI classes × 2 omics categories × 10 markers; Fig. 3d), 216 (2 BMI classes × 4 omics categories × 27 features; Fig. 3e), 24 (2 BMI classes × 4 omics categories × 3 metrics; Fig. 4b), or 24 (2 BMI classes × 12 measures; Supplementary Fig. 6c) regressions. In the above regression analyses for the TwinsUK cohort, ancestry PCs were eliminated from the covariates due to data availability.

## Supporting information

TRIPOD checklist

## Data Availability

The de-identified Arivale datasets that were used in this study can be accessed by qualified researchers for research purposes. Requests should be sent to, and the data will be available after submission and approval of a research plan. The de-identified TwinsUK datasets that were used in this study were provided by Department of Twin Research & Genetic Epidemiology
(King's College London) after the approval of our Data Access Application (Project Number: E1192).
Requests should be referred to their website (http://twinsuk.ac.uk/resources-for-researchers/access-our-data/).

## Data visualization

Results were visualized using Python matplotlib (version 3.4.3) and seaborn (version 0.11.2) libraries, except for the plasma analyte correlation network. Data were summarized as the mean with 95% confidence interval (CI) or the boxplot (median: center line; 95% CI around median: notch; [*Q*_1_, *Q*_3_]: box limits; [*x*_min_, *x*_max_]: whiskers, where *Q*_1_ and *Q*_3_ are the 1st and 3rd quartile values, and *x*_min_ and *x*_max_ are the minimum and maximum values in [*Q*_1_ − 1.5 × IQR, *Q*_3_ + 1.5 × IQR] (IQR: the interquartile range, *Q*_3_ − *Q*_1_), respectively), as indicated in each figure legend. For presentation purpose, CI was simultaneously calculated during visualization using Python seaborn *barplot* or *boxplot* API with default setting (1,000 times bootstrapping or a Gaussian-based asymptotic approximation, respectively). The OLS linear regression line with 95% CI was simultaneously generated during visualization using Python seaborn *regplot* API with default setting (1,000 times bootstrapping). The plasma analyte correlation network was visualized with a circos plot using R circlize (version 0.4.15) package^70^.

## Data availability

The de-identified Arivale datasets that were used in this study can be accessed by qualified researchers for research purposes. Requests should be sent to, and the data will be available after submission and approval of a research plan. The de-identified TwinsUK datasets that were used in this study were provided by Department of Twin Research & Genetic Epidemiology (King’s College London) after the approval of our Data Access Application (Project Number: E1192).

Requests should be referred to their website (http://twinsuk.ac.uk/resources-for-researchers/access-our-data/).

## Code availability

Code used in this study is freely available on GitHub (https://github.com/PriceLab/Multiomics-BMI).

## Acknowledgements

We thank Sergey A. Kornilov, Gustavo Glusman, and Max Robinson (Institute for Systems Biology; ISB) for providing comments to this study. We thank Victoria Vazquez and Andrew Anastasiou (King’s College London) for their support in obtaining and utilizing the TwinsUK data access. We are grateful to all Arivale and TwinsUK participants who consented to using their deidentified data for research purposes. This work was supported by the M.J. Murdock Charitable Trust (Reference No. 2014096:MNL:11/20/2014, awarded to N.D.P. and L.H.), the National Institutes of Health (NIH) grants awarded by the National Institute on Aging (NIA) (U19AG023122 and 5U01AG061359), and a generous gift from K. Carole Ellison (to K.W., T.W., and A.Z.). K.W. was supported by The Uehara Memorial Foundation (Overseas Postdoctoral Fellowships). C.D. and S.M.G. were supported by the Washington Research Foundation Distinguished Investigator Award and startup funds from ISB. TwinsUK is funded by the Wellcome Trust, Medical Research Council, Versus Arthritis, European Union Horizon 2020, Chronic Disease Research Foundation (CDRF), Zoe Ltd, the National Institute for Health and Care Research (NIHR) Clinical Research Network (CRN) and Biomedical Research Centre based at Guy’s and St Thomas’ NHS Foundation Trust in partnership with King’s College London.

## Author Contribution

K.W., T.W., L.H., N.D.P., and N.R. conceptualized the study. K.W., T.W., A.Z., N.D.P., and N.R. participated in the study design. K.W., T.W., C.D., B.L., and N.R. performed data analysis and figure generation. C.D., J.C.E., J.J.H., J.C.L., S.M.G., A.T.M., and L.H. assisted in results interpretation.

J.C.L. and A.T.M. managed the logistics of data collection and integration. K.W., T.W., and N.R. were the primary authors of the paper, with contributions from all other authors. All authors read and approved the final manuscript.

## Competing Interests

J.J.H. has received grants from Pfizer and Novartis for research unrelated to this study. All other authors declare no competing interests.

## Notes

### Summary of Updates

The main change in this version is the addition of the TwinsUK validation dataset.

