## Supplementary Information for "Multiomic Body Mass Index signatures in blood reveal clinically relevant population heterogeneity and variable responses to a healthy lifestyle intervention"

**This PDF file includes:**

Supplementary Figures 1 to 8
Table legends for Supplementary Data 1 to 10

**Other Supplementary Information files in this study include the following:**

Supplementary Data 1 to 10
TRIPOD Checklist

### Supplementary Figures

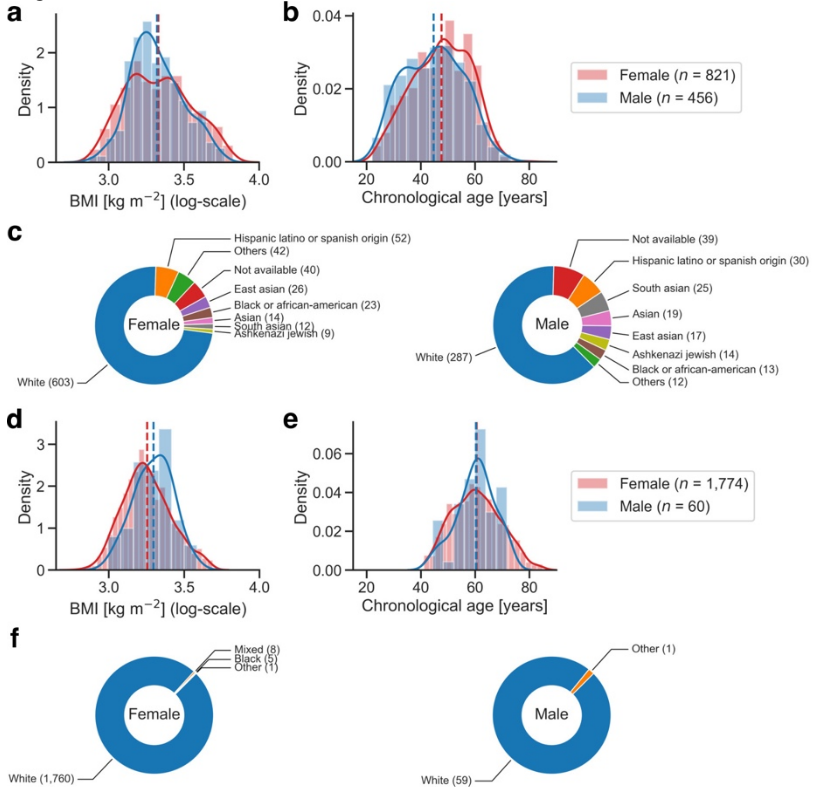


**Supplementary Figure 1. Demographic information of study cohorts.**

**a**–**c** Demographic information of the Arivale study cohort (Fig. 1a, *n* = 1,277 participants). **d**–**f** Demographic information of the TwinsUK study cohort (Fig. 1a, *n* = 1,834 participants). **a**, **b**, **d**, **e** Distribution of the baseline Body Mass Index (BMI) (**a**, **d**) or age (**b**, **e**). *n* = 821 (**a**, **b**; Female), 456 (**a**, **b**; Male), 1,774 (**d**, **e**; Female), 60 (**d**, **e**; Male) participants. The solid and dashed lines indicate the kernel density estimate and the mean of BMI (**a**, Female: 28.6 kg $m^{-2}$; **a**, Male: 28.1 kg $m^{-2}$; **d**, Female: 26.2 kg $m^{-2}$; **d**, Male: 27.1 kg $m^{-2}$) or age (**b**, Female: 47.6 years; **b**, Male: 44.7 years; **e**, Female: 61.4 years; **e**, Male: 62.0 years), respectively. **c**, **f** Composition of self-reported race (**c**) or ethnicity (**f**). The number in parentheses indicates the number of participants.

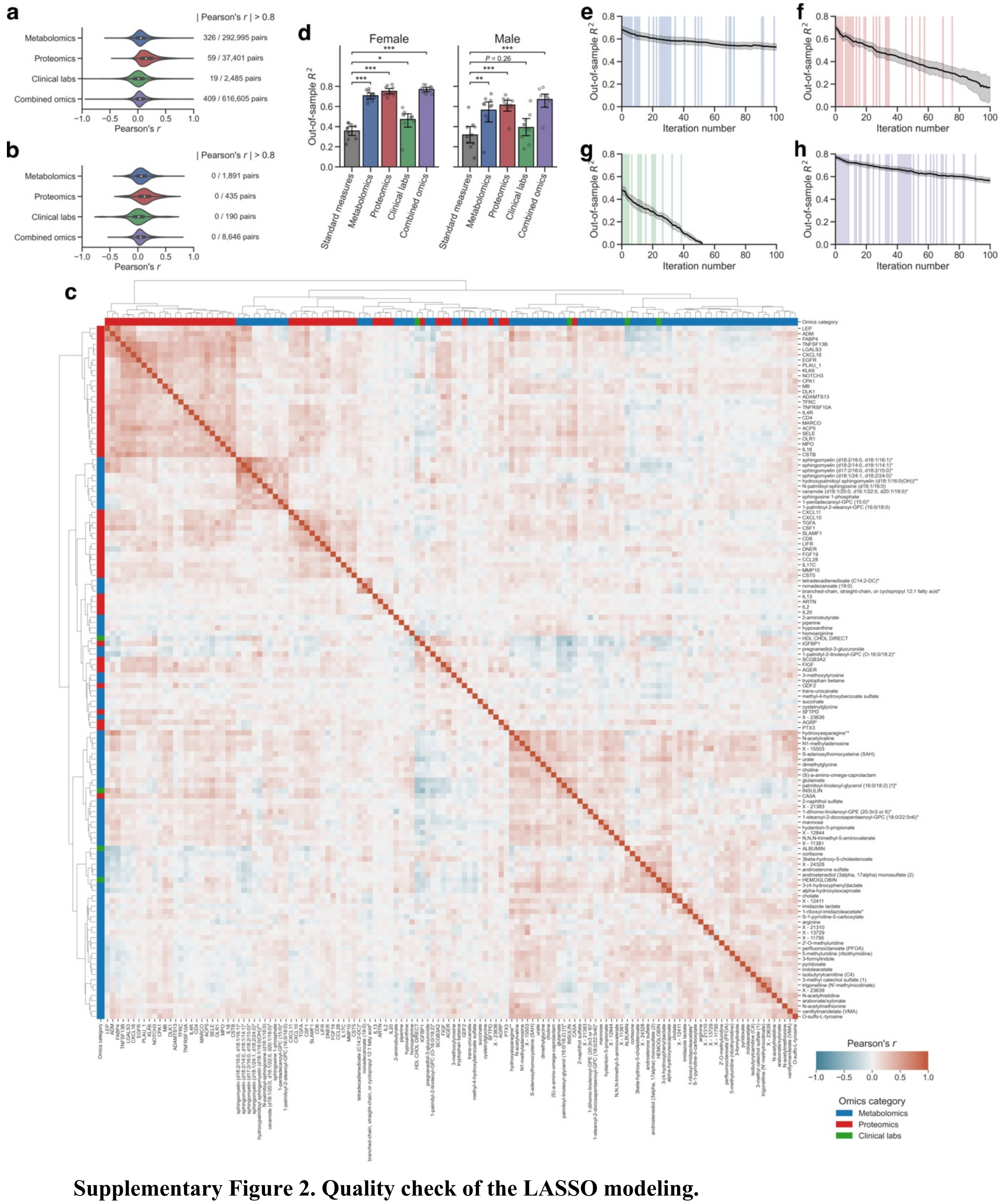


**Supplementary Figure 2. Quality check of the LASSO modeling.**

**a**, **b** Pairwise correlation of all plasma analytes (**a**; Metabolomics: 766 metabolites, Proteomics: 274 proteins, Clinical labs: 71 clinical laboratory tests, Combined omics: 1,111 analytes) or the analytes that were retained across all ten least absolute shrinkage and selection operator (LASSO) models (**b**; Metabolomics: 62 metabolites, Proteomics: 30 proteins, Clinical labs: 20 clinical laboratory tests, Combined omics: 132 analytes). Each violin is scaled to have same width between the omics categories and represents the kernel density distribution with boxplot (median: white point, [$Q_1$, $Q_3$]: box limits, [$x_{min}$, $x_{max}$]: whiskers, where $Q_1$ and $Q_3$ are the 1st and 3rd quartile values, and $x_{min}$ and $x_{max}$

are the minimum and maximum values in [$Q_1 - 1.5 \times$ IQR, $Q_3 + 1.5 \times$ IQR] (IQR: the interquartile range, $Q_3 - Q_1$), respectively). **c** Hierarchical clustering and heatmap for the pairwise correlations of the analytes that were retained across all ten combined omics-based Body Mass Index (BMI) models (132 analytes: 77 metabolites, 51 proteins, 4 clinical laboratory tests). Of note, both upper and lower triangular sides of the symmetric matrix are visualized. **d** Model performance of each fitted BMI model with sex stratification. Out-of-sample $R^2$ was calculated from each corresponding hold-out testing set. Standard measures: ordinary least squares (OLS) linear regression model with sex, age, triglycerides, high-density lipoprotein (HDL)-cholesterol, low-density lipoprotein (LDL)-cholesterol, glucose, insulin, and homeostatic model assessment for insulin resistance (HOMA-IR) as regressors. Data: mean with 95% confidence interval (CI), $n = 10$ models. *Adjusted $P < 0.05$, **adjusted $P < 0.01$, ***adjusted $P < 0.001$ in two-sided Welch's $t$-test with the Benjamini–Hochberg method across the eight (four comparisons × 2 sexes) comparisons. Note that the sample size for modeling was different between female and male (Female: 821 participants, Male: 456 participants). **e**–**h** Transition of out-of-sample $R^2$ in the LASSO-modeling iteration analysis for metabolomics (**e**), proteomics (**f**), clinical labs (**g**), or combined omics (**h**). At the end of each iteration, the variable that was retained across ten models and that had the highest absolute value for the mean of ten $\beta$-coefficients was removed from the input omic dataset. The iteration is highlighted with shading color when the removed analyte is the variable that was retained across all the original ten models. Data: mean with 95% CI, $n = 10$ models.

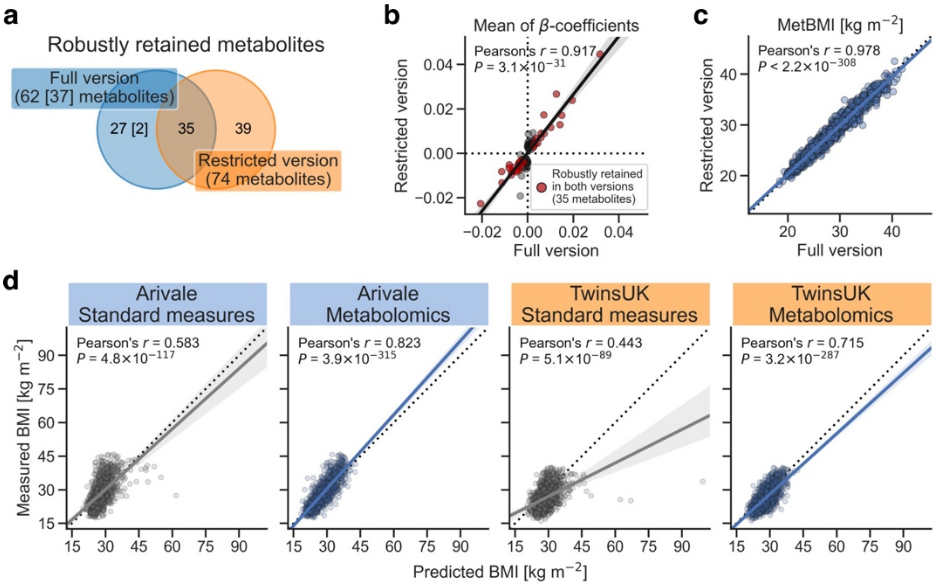


**Supplementary Figure 3. The restricted metabolomics-based BMI model predominantly maintained the characteristics of the original full model.**

**a**–**c** Comparison of the metabolomics-based Body Mass Index (MetBMI) model between the main analyses (Arivale cohort) and the validation analyses (TwinsUK cohort). Full version: least absolute shrinkage and selection operator (LASSO) model trained by all 766 metabolites in the Arivale dataset, Restricted version: LASSO model trained by the common 489 metabolites in the Arivale and TwinsUK datasets. **a** The number of the variables that were robustly retained across all ten MetBMI models. The number in square brackets indicates the number of the robustly retained metabolites that were derived from the common 489 metabolites. **b** Correlation of the mean of $\beta$-coefficients in the ten MetBMI models. Only the robustly retained metabolites in either full version (37 metabolites) or restricted version (74 metabolites) were analyzed. **c** Correlation of the MetBMI prediction. **b**, **c** The solid line is the ordinary least squares (OLS) linear regression line with 95% confidence interval (CI), and the dotted line in **c** is the value in full version = the value in restricted version. *P*: *P*-value of two-sided Pearson's correlation test. $n = 76$ metabolites (**b**), 1,277 participants (**c**). **d** Correlation between the measured and predicted BMIs. The solid line is the OLS linear regression line with 95% CI, and the dotted line is measured BMI = predicted BMI. Standard measures: OLS linear regression model with sex, age, triglycerides, high-density lipoprotein (HDL)-cholesterol, low-density lipoprotein (LDL)-cholesterol, glucose, insulin, and homeostatic model assessment for insulin resistance (HOMA-IR) as regressors; Metabolomics: the restricted version of MetBMI model, corresponding to Metabolomics (restricted) in Fig. 1d; *P*: adjusted *P*-value of two-sided Pearson's correlation test with the Benjamini–Hochberg method across the four (two categories × two cohorts) tests. $n = 1,277$ (Arivale), 1,834 (TwinsUK) participants.

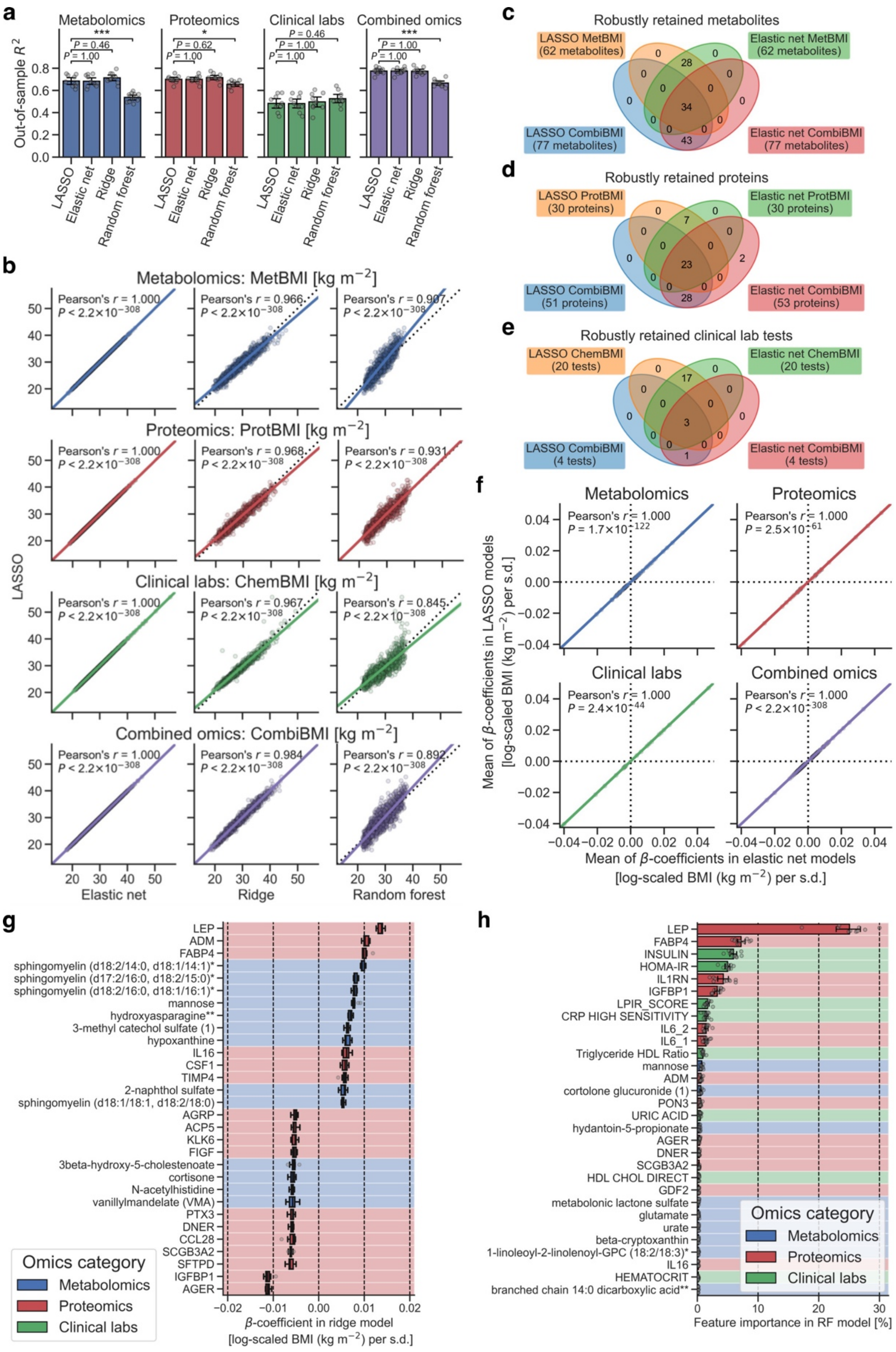


**Supplementary Figure 4. Omics-based BMI models were similar between LASSO and the other methods.**

**a** Model performance of each fitted Body Mass Index (BMI) model. Out-of-sample $R^2$ was calculated from each corresponding hold-out testing set. Data: mean with 95% confidence interval (CI), $n = 10$ models. *Adjusted $P < 0.05$, ***Adjusted $P < 0.001$ in two-sided Welch's $t$-test with the Benjamini–Hochberg method across the 12 (3 methods × 4 categories) comparisons. **b** Correlation of the predicted BMI between LASSO and the other methods. The solid line is the ordinary least squares (OLS) linear regression line with 95% CI, and the dotted line is the value in LASSO = the value in the other method. *P*: adjusted *P*-value of two-sided Pearson's correlation test with the Benjamini–Hochberg method across the 12 (3 methods × 4 categories) combinations. $n = 1{,}277$ participants. **c–f** Comparison of the omics-based BMI model between LASSO and elastic net (EN) methods. **c–e** The number of the variables that were robustly retained across all ten LASSO or EN models. MetBMI: metabolomics-based BMI model, ProtBMI: proteomics-based BMI model, ChemBMI: clinical chemistries-based BMI model, CombiBMI: combined omics-based BMI model. **f** Correlation of the mean of $\beta$-coefficients in the ten omics-based BMI models. Only the robustly retained analytes in either LASSO models or EN models were analyzed. The solid line is the OLS linear regression line with 95% CI. *P*: adjusted *P*-value of two-sided Pearson's correlation test with the Benjamini–Hochberg method across the four categories. $n = 62$ metabolites (Metabolomics), 30 proteins (Proteomics), 20 clinical laboratory tests (Clinical labs), 134 analytes (Combined omics). **g** The top 30 variables that had the highest absolute value for the mean of $\beta$-coefficients in the ten ridge CombiBMI models. $\beta$-coefficient was obtained from the fitted CombiBMI model with ridge regression. Data: median (center line), $[Q_1, Q_3]$ (box limits), $[x_{min}, x_{max}]$ (whiskers), where $Q_1$ and $Q_3$ are the 1st and 3rd quartile values, and $x_{min}$ and $x_{max}$ are the minimum and maximum values in $[Q_1 - 1.5 \times \text{IQR}, Q_3 + 1.5 \times \text{IQR}]$ (IQR: the interquartile range, $Q_3 - Q_1$), respectively; $n = 10$ models. **h** The top 30 variables that had the highest mean of feature importance in the ten random forest (RF) CombiBMI models. The importance of a feature was calculated as the normalized total reduction of the mean squared error that was brought by the feature. Data: mean with 95% CI, $n = 10$ models. **g**, **h** Each background color corresponds to the analyte category.

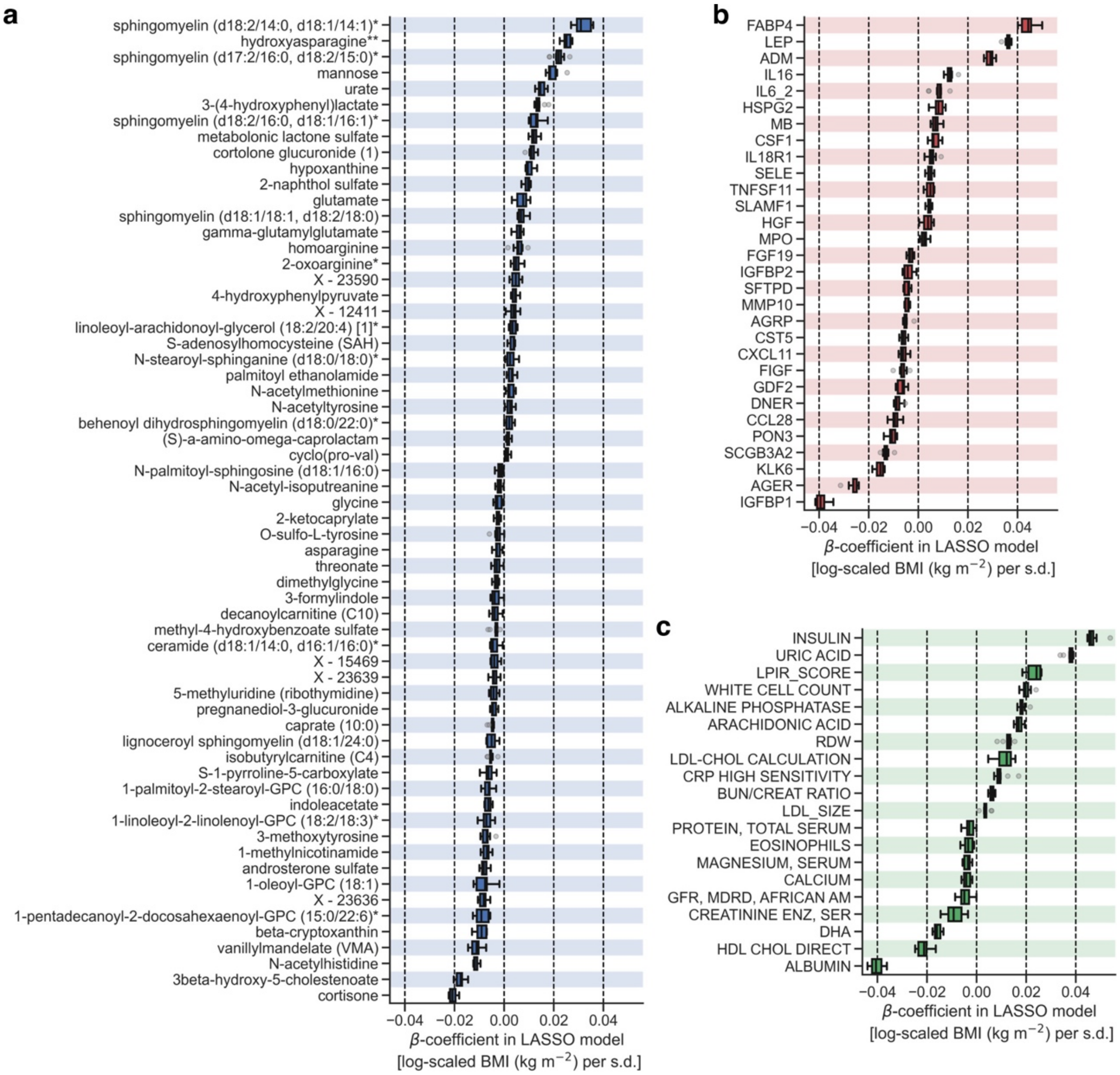


**Supplementary Figure 5. Variable diversity and contribution to the omics-based BMI model were different between omics categories.**

**a**–**c** The variables that were retained across all ten metabolomics-based (**a**), proteomics-based (**b**), or clinical labs-based (**c**) Body Mass Index (BMI) models (**a**: 62 metabolites, **b**: 30 proteins, **c**: 20 clinical laboratory tests). $\beta$-coefficient was obtained from the fitted omics-based BMI model with least absolute shrinkage and selection operator (LASSO) regression. Data: median (center line), [$Q_1$, $Q_3$] (box limits), [$x_{\min}$, $x_{\max}$] (whiskers), where $Q_1$ and $Q_3$ are the 1st and 3rd quartile values, and $x_{\min}$ and $x_{\max}$ are the minimum and maximum values in [$Q_1 - 1.5 \times \text{IQR}$, $Q_3 + 1.5 \times \text{IQR}$] (IQR: the interquartile range, $Q_3 - Q_1$), respectively; $n = 10$ models.

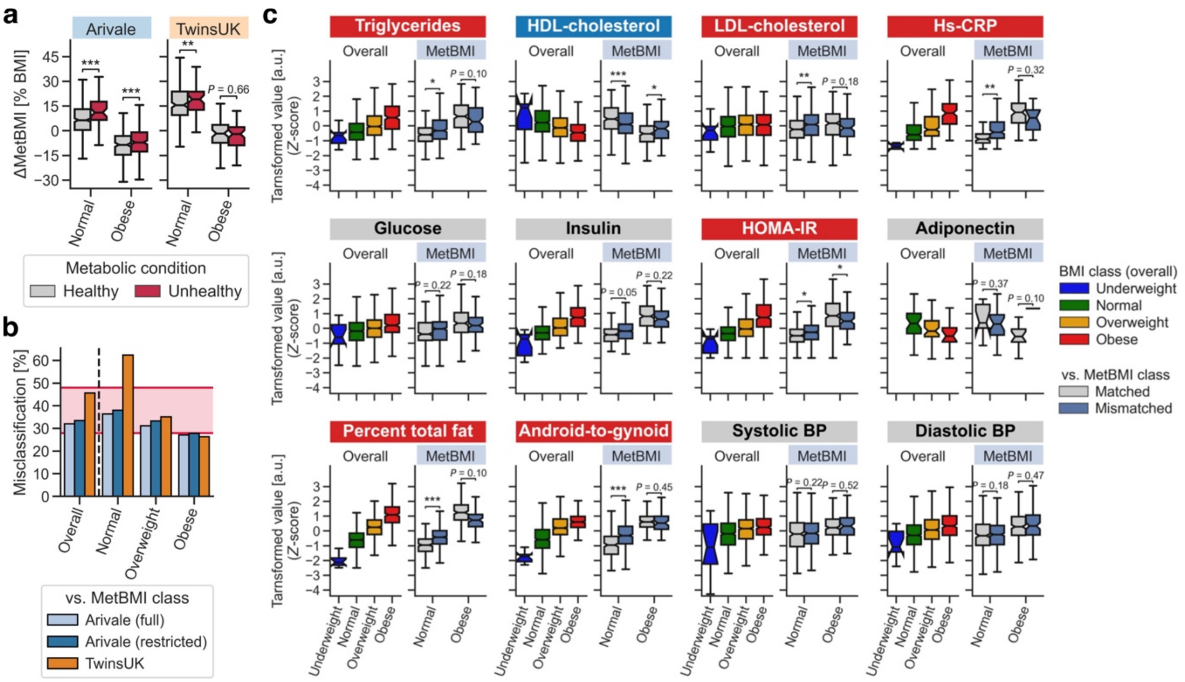


**Supplementary Figure 6. The metabolic heterogeneity within the standard BMI classes was validated with the TwinsUK cohort.**

**a** Difference in ΔMetBMI (i.e., difference of the metabolomics-inferred Body Mass Index (MetBMI) from the measured BMI) between clinically-defined metabolic health conditions. Significance was assessed using ordinary least squares (OLS) linear regression with BMI, sex, and age as covariates, while adjusting multiple testing with the Benjamini–Hochberg method across the four (two BMI classes × two cohorts) regressions. For Arivale cohort, ancestry principal components (PCs) were also included in the covariates. MetBMI in Arivale was derived from the MetBMI model trained by the common 489 metabolites in the Arivale and TwinsUK datasets, corresponding to the restricted version in Supplementary Fig. 3. **b** Misclassification rate of overall cohort or each BMI class against MetBMI class. Arivale (full): based on the full version of MetBMI model in Supplementary Fig. 3 (i.e., the same with the corresponding ones in Fig. 3c), Arivale (restricted): based on the restricted version of MetBMI model in Supplementary Fig. 3. Range of the previously reported misclassification rate is highlighted with pink background. Note that the underweight BMI class is not presented due to small sample size, but its misclassification rate was 100% against all omics-based BMI classes. **c** Difference in the obesity-related phenotypic measure between Matched and Mismatched groups in the TwinsUK cohort. Significance was assessed using OLS linear regression with BMI, sex, and age as covariates, while adjusting multiple testing with the Benjamini–Hochberg method across the 24 (2 BMI classes × 12 measures) regressions. HDL: high-density lipoprotein, LDL: low-density lipoprotein, Hs-CRP: high-sensitivity C-reactive protein, Percent total fat: percentage of total fat in whole body, Android-to-gynoid: ratio of fat in android region to fat in gynoid region, HOMA-IR: homeostatic model assessment for insulin resistance, BP: blood pressure. **a**, **c** Data: median (center line), 95% confidence interval (CI) around median (notch), $[Q_1, Q_3]$ (box limits), $[x_{min}, x_{max}]$ (whiskers), where $Q_1$ and $Q_3$ are the 1st and 3rd quartile values, and $x_{min}$ and $x_{max}$ are the minimum and maximum values in $[Q_1 - 1.5 \times \text{IQR}, Q_3 + 1.5 \times \text{IQR}]$ (IQR: the interquartile range, $Q_3 - Q_1$), respectively; $n = 373$ (**a**, Healthy in Normal of Arivale), 49 (**a**, Unhealthy in Normal of Arivale), 208 (**a**, Healthy in Obese of Arivale), 241 (**a**, Unhealthy in Obese of Arivale), 209 (**a**, Healthy in Normal of TwinsUK), 50 (**a**, Unhealthy in Normal of TwinsUK), 64 (**a**, Healthy in Obese of TwinsUK), 57 (**a**, Unhealthy in Obese of TwinsUK) participants (see Supplementary Data 6 for each sample size in **c**). *Adjusted $P < 0.05$, **adjusted $P < 0.01$, ***adjusted $P < 0.001$.

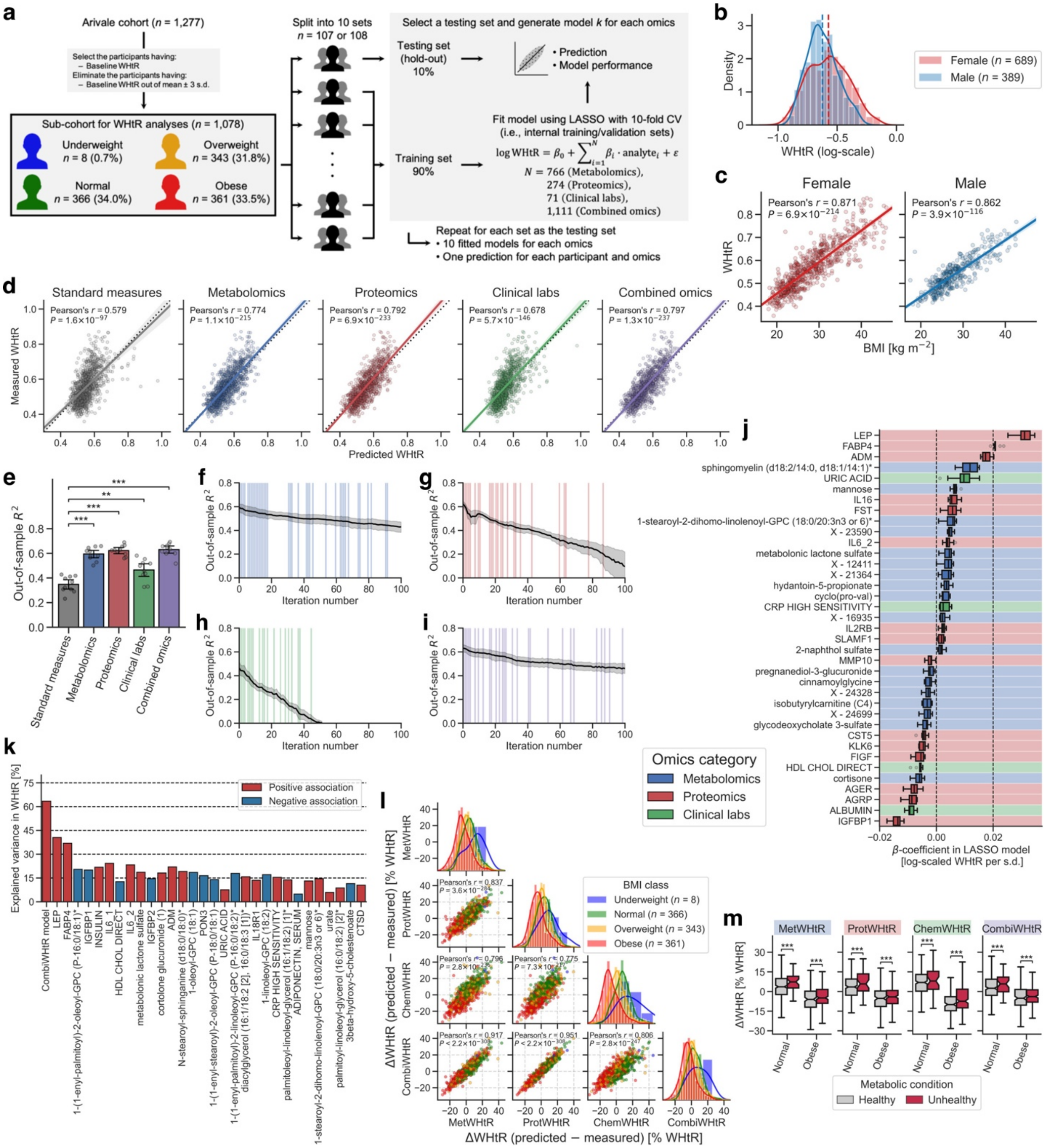


**Supplementary Figure 7. Omics-based WHtR models consistently supported the findings of omics-based BMI models.**

**a** Overview of study cohort and the omics-based waist-to-height ratio (WHtR) model generation. LASSO: least absolute shrinkage and selection operator, CV: cross-validation. **b** Distribution of the baseline WHtR. *n* = 689 (Female), 389 (Male) participants. The solid and dashed lines indicate the kernel density estimate and the mean of WHtR (Female: 0.571, Male: 0.539 [raw scale]), respectively. **c** Correlation between the measured WHtR and Body Mass Index (BMI). The solid line is the ordinary least squares (OLS) linear regression line with 95% confidence interval (CI). *P*: adjusted *P*-value of two-sided Pearson's correlation test with the Benjamini–Hochberg method across the two sexes. *n* = 689 (Female), 389 (Male) participants. **d** Correlation between the measured and predicted WHtRs.

The solid line is the OLS linear regression line with 95% CI, and the dotted line is measured WHtR = predicted WHtR. Standard measures: OLS linear regression model with sex, age, triglycerides, high-density lipoprotein (HDL)-cholesterol, low-density lipoprotein (LDL)-cholesterol, glucose, insulin, and homeostatic model assessment for insulin resistance (HOMA-IR) as regressors; *P*: adjusted *P*-value of two-sided Pearson's correlation test with the Benjamini–Hochberg method across the five categories. $n$ = 1,078 participants. **e** Model performance of each fitted WHtR model. Out-of-sample $R^2$ was calculated from each corresponding hold-out testing set. Data: mean with 95% CI, $n$ = 10 models. **Adjusted $P < 0.01$, adjusted $P < 0.001$ in two-sided Welch's *t*-test with the Benjamini–Hochberg method across the four comparisons. **f**–**i** Transition of out-of-sample $R^2$ in the LASSO-modeling iteration analysis for metabolomics (**f**), proteomics (**g**), clinical labs (**h**), or combined omics (**i**). At the end of each iteration, the variable that was retained across ten models and that had the highest absolute value for the mean of ten $\beta$-coefficients was removed from the input omic dataset. The iteration is highlighted with shading color when the removed analyte is the variable that was retained across all the original ten models. Data: mean with 95% CI, $n$ = 10 models. **j** The variables that were retained across all ten combined omics-based WHtR (CombiWHtR) models (37 analytes: 18 metabolites, 15 proteins, and 4 clinical laboratory tests). $\beta$-coefficient was obtained from the fitted CombiWHtR model with LASSO regression. Each background color corresponds to the analyte category. Data: median (center line), [$Q_1$, $Q_3$] (box limits), [$x_{min}$, $x_{max}$] (whiskers), where $Q_1$ and $Q_3$ are the 1st and 3rd quartile values, and $x_{min}$ and $x_{max}$ are the minimum and maximum values in [$Q_1 - 1.5 \times$ IQR, $Q_3 + 1.5 \times$ IQR] (IQR: the interquartile range, $Q_3 - Q_1$), respectively; $n$ = 10 models. **k** Univariate explained variance in WHtR by each analyte. WHtR was independently regressed on each of the analytes that were retained in at least one of the ten CombiWHtR models (288 analytes; Supplementary Data 9), using OLS linear regression with sex, age, and ancestry principal components (PCs) as covariates. Multiple testing was adjusted with the Benjamini–Hochberg method across the 289 regressions, including CombiWHtR model as reference. Among the analytes that were significantly associated with WHtR (212 analytes), only the top 30 significant analytes are presented with their univariate variances. **l** Difference of the omics-inferred WHtR from the measured WHtR (ΔWHtR). MetWHtR: metabolomics-inferred WHtR, ProtWHtR: proteomics-inferred WHtR, ChemWHtR: clinical chemistries-inferred WHtR, CombiWHtR: combined omics-inferred WHtR, *P*: adjusted *P*-value of two-sided Pearson's correlation test with the Benjamini–Hochberg method across the six combinations, $n$: the number of participants in each BMI class (total $n$ = 1,078 participants). The line in histogram panels indicates the kernel density estimate. **m** Difference in ΔWHtR between clinically-defined metabolic health conditions. Significance was assessed using OLS linear regression with WHtR, sex, age, and ancestry PCs as covariates, while adjusting multiple testing with the Benjamini–Hochberg method across the eight (two BMI classes × four omics categories) regressions. Data: each boxplot metric is the same with **j**, with the addition of 95% CI around median (notch); $n$ = 320 (Healthy in Normal), 42 (Unhealthy in Normal), 164 (Healthy in Obese), 197 (Unhealthy in Obese) participants. ***Adjusted $P < 0.001$.

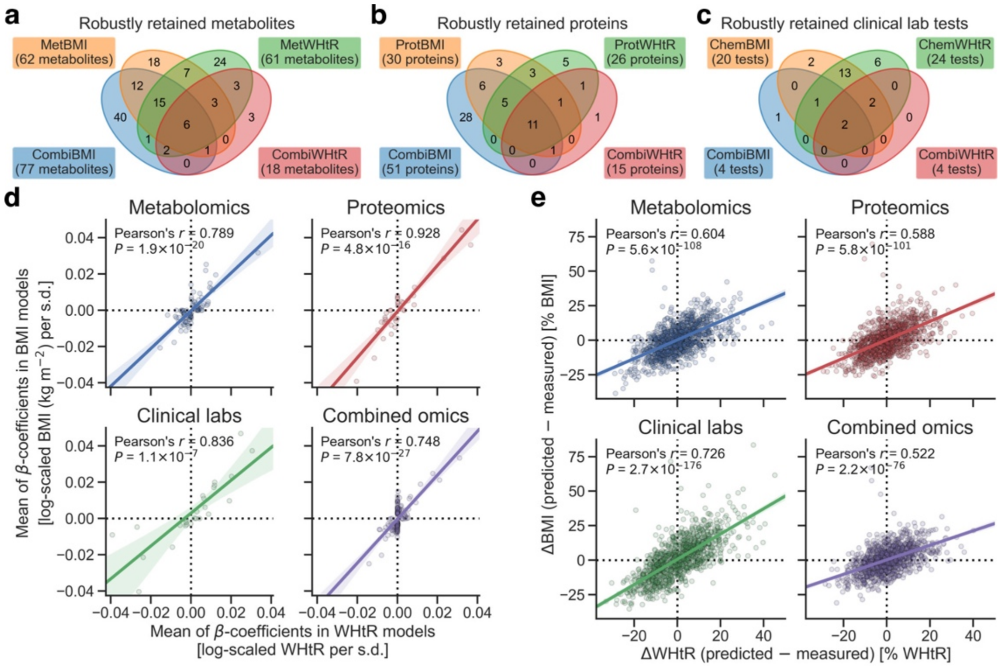


**Supplementary Figure 8. Predominant commonality with minor specificity was observed between the omics-based BMI and WHtR models.**

**a**–**d** Comparison of the omics-based least absolute shrinkage and selection operator (LASSO) model between Body Mass Index (BMI) and waist-to-height ratio (WHtR). **a**–**c** The number of the variables that were robustly retained across all ten LASSO models. MetBMI: metabolomics-based BMI model, MetWHtR: metabolomics-based WHtR model, ProtBMI: proteomics-based BMI model, ProtWHtR: proteomics-based WHtR model, ChemBMI: clinical chemistries-based BMI model, ChemWHtR: clinical chemistries-based WHtR model, CombiBMI: combined omics-based BMI model, CombiWHtR: combined omics-based WHtR model. **d** Correlation of the mean of *β*-coefficients in the ten LASSO models. Only the robustly retained analytes in either BMI models or WHtR models were analyzed. **e** Correlation between ΔBMI (i.e., difference of the omics-inferred BMI from the measured BMI) and ΔWHtR (i.e., difference of the omics-inferred WHtR from the measured WHtR). Only the participants having both BMI and WHtR were analyzed. **d**, **e** The solid line is the ordinary least squares (OLS) linear regression line with 95% confidence interval (CI). *P*: adjusted *P*-value of two-sided Pearson's correlation test with the Benjamini–Hochberg method across the four categories. $n$ = 92 metabolites (**d**, Metabolomics), 36 proteins (**d**, Proteomics), 26 clinical laboratory tests (**d**, Clinical labs), 146 analytes (**d**, Combined omics), 1,078 participants (**e**).

### Table legends for Supplementary Data

**Supplementary Data 1. Cohort summary.**

This .xlsx file contains demographic summary of the study cohorts and statistical test summaries for the independency of split sets. Descriptions about each sheet and each column are included in the README sheet.

**Supplementary Data 2. Analytes of blood-measured omics.**

This .xlsx file contains information about the analytes of blood-measured omics and basic statistics of their baseline measurements. Descriptions about each sheet and each column are included in the README sheet.

**Supplementary Data 3. *β*-coefficient estimates for the variables of the omics-based BMI models.**

This .xlsx file contains *β*-coefficient estimates for the variables of the omics-based BMI models, related to Figure 2 and Supplementary Figure 2–5, 8. Descriptions about each sheet and each column are included in the README sheet.

**Supplementary Data 4. Relationships of the numeric physiological measures with the measured or omics-inferred BMI.**

This .xlsx file contains the regression analysis summary for the association between each of the 51 numeric physiological measures and the measured or omics-inferred BMI, corresponding to Figure 1e. Descriptions about each column are included in the README sheet.

**Supplementary Data 5. Relationships of the retained analytes in the omics-based BMI models with BMI.**

This .xlsx file contains the regression analysis summary for the association between BMI and each of the analytes that were retained in at least one of ten LASSO models, corresponding to Figure 2b–d. Descriptions about each sheet and each column are included in the README sheet.

**Supplementary Data 6. Differences in phenotypic measures between the misclassification strata against the omics-inferred BMI class.**

This .xlsx file contains the regression analysis summary for the difference in the obesity-related clinical blood marker, the BMI-associated numeric physiological feature, or the gut microbiome α-diversity metric between the misclassification strata against the omics-inferred BMI class, corresponding to Figure 3d, 3e, 4b and Supplementary Figure 6c. Descriptions about each sheet and each column are included in the README sheet.

**Supplementary Data 7. Plasma analyte correlations modified by the baseline metabolic state and by lifestyle intervention.**

This .xlsx file contains the interaction analysis summary for the plasma analyte correlations modified by the baseline MetBMI and by days in program, corresponding to Figure 6. Descriptions about each column are included in the README sheet.

**Supplementary Data 8. *β*-coefficient estimates for the variables of the omics-based WHtR models.**

This .xlsx file contains $\beta$-coefficient estimates for the variables of the omics-based WHtR models, related to Supplementary Figure 7, 8. Descriptions about each sheet and each column are included in the README sheet.

**Supplementary Data 9. Relationships of the retained analytes in the omics-based WHtR models with WHtR.**

This .xlsx file contains the regression analysis summary for the association between WHtR and each of the analytes that were retained in at least one of ten LASSO models, corresponding to Supplementary Figure 7k. Descriptions about each sheet and each column are included in the README sheet.

**Supplementary Data 10. Statistical test summary.**

This .xlsx file contains the statistical test summary including sample size, degrees of freedom, test statistic, (nominal) *P*-value, and adjusted *P*-value, corresponding to Figure 1b–d, 3a, 3b, 4c–f and Supplementary Figure 2d, 3d, 4a, 4b, 4f, 6a, 7c–e, 7l, 7m, 8d, 8e. Descriptions about each sheet and each column are included in the README sheet.
